# Informing risk stratification and vaccine policy via a predictive mathematical model of SARS-COV-2 infection

**DOI:** 10.64898/2026.10.12.26365315

**Authors:** Benny Chain, Curtis G. Callan, David Scobie, Matthew V. Cowley, Jana Jiang, Rishi K. Gupta, Carolin T. Turner, Joshua Rosenheim, Clare Thakker, Lucy Bell, Gayathri Nageswaran, Suzanne Byrne, Ben Killingley, Mariya Kalinova, Alison Boyers, Andrew Catchpole, Alex Mann, Rik G. H. Lindeboom, Marko Z. Nikolic, Sarah A. Teichmann, Helen Wagstaffe, Andreas Tiffeau-Mayer, Christopher Chiu, Mahdad Noursadeghi

**Affiliations:** Institute of Infection, Immunity and Transplantation, University College London; London, UK; Department of Computer Science; University College London; London, UK; Department of Physics, Princeton University; Princeton NJ, USA; Département de Physique, École Normale Supérieure; Paris, France; Division of Infection, University College London Hospital; London, UK; hVIVO; London, UK; Wellcome Sanger Institute, Wellcome Genome Campus; Cambridge, UK; The Netherlands Cancer Institute; Amsterdam, NL; UCL Respiratory, Division of Medicine, University College London; London, UK; Cambridge Stem Cell Institute, Jeffrey Cheah Biomedical Centre, Cambridge Biomedical Campus, University of Cambridge; Cambridge, UK; Department of Medicine, University of Cambridge; Cambridge, UK; CIFAR Macmillan Multi-scale Human Programme, CIFAR; Toronto, Canada; Department of Infectious Disease, Imperial College London; London, UK; Institute for the Physics of Living Systems, University College London, London, UK

## Abstract

SARS-CoV-2 human challenge studies enabled unprecedented high frequency temporal measurements of immune determinants of viral control. We integrated prior knowledge of SARS-CoV-2 replication, innate, cytotoxic T cell and antibody responses into a dynamic model which captured viral-load kinetics in seronegative volunteers challenged with wildtype virus. The model predicted measured innate and adaptive immune dynamics. Perturbation analyses indicated that innate immunity limits peak viral load, whereas antibodies principally mediate clearance while cytotoxic T cells become critical in antibody deficiency. Pre-existing antibodies suppressed both viral replication and subsequent immune activation- a prediction validated in a second challenge study of seropositive participants infected with the SARS-CoV-2 Delta variant. The model explains how specific immunodeficiencies may increase susceptibility to severe or persistent infection, supporting clinical risk stratification. It suggests that T cell-targeted vaccination strategies may provide limited protection without effective neutralising antibodies, and that attenuated boosting after low-dose exposure requires repeated vaccination to reduce breakthrough infections, informing public health policy.

**One Sentence Summary:** A mathematical model of SARS-COV-2 infection predicts innate and adaptive immunity, and informs risk stratification and vaccine policy.

## INTRODUCTION

SARS-CoV-2 caused one of the largest recorded human pandemics since the 1918 influenza pandemic, spreading rapidly across the globe and infecting most of the world population. In immunocompetent individuals viral transmission usually occurs in a short window early in the course of infection (*1–4*) and a quantitative understanding of the dynamics of in-host/virus interactions during this period is therefore important for understanding between-host spread and designing effective therapeutic interventions. However, observational studies of SARS-CoV-2 infection usually miss the earliest phase of the infection, since they are dependent on the detection of symptoms. Longitudinal studies of at risk populations during periods of high rates of infection were able to infer some features of the early immune response (*5, 6*), but quantitative analysis was limited because of logistic limitations to sampling frequency and uncertainty about the time of infection onset.

In situations where data collection is challenging, and experimental perturbation limited by ethical and logistical factors, mathematical modelling has been useful in probing latent features of the host/virus interaction, which may be difficult or impossible to observe directly. The application of mathematical modelling to virus infections has a long well-established track record (*7*). Modelling, for example, provided key evidence for the existence of the long latent period of HIV infection, which was hard to measure experimentally (*8, 9*). Several mathematical models of in-host SARS-Cov-2 infection have also been published (reviewed in (*10*)).

In this study we integrate existing knowledge of the human immune response to viruses to formulate a mechanistic mathematical model, which explicitly incorporates three strands of immunity, innate anti-viral immunity (of which a key element is the Type I interferon response), cytotoxic T cell immunity, and antibody-dependent viral neutralisation. All three effector mechanisms have been implicated in SARS-Cov-2 infection (*5, 11–14*). We build the model using only well-established fundamental prior knowledge of the human immune system, and tune the model parameters by fitting to densely sampled viral load measurements obtained during a healthy volunteer challenge study (*15*). This study was carried out early in the pandemic using the original pre-alpha wildtype strain of SARS-CoV-2 and was motivated by the need for a platform for rapid testing of novel vaccines and therapeutics. The study provided a rare opportunity to access human samples at the earliest stages of acute viral infection, and has already provided rich, multimodal and highly-sampled data that have led to novel perspectives on the earliest events of SARS-CoV-2 host interaction (*16–19*). After parametrising the model based only on viral load dynamics, we explore how the predictions of the model fit the observed immunological data from the challenge study, including unpublished longitudinal blood global and T cell receptor (TCR) transcriptomics. We perturb the model and explore its predictions of virus and host immune dynamics under conditions of compromised immunity, or of pre-existing immunity. We examine these predictions in the context of published data on immune-compromised individuals, and new unpublished transcriptomic data from a further challenge study of sero-positive individuals infected with the delta strain of SARS-CoV-2(*20*). The agreement between model and observations under a range of perturbation suggest that the model captures some of the fundamental features of the host response to SARS-CoV-2. The model adds a quantitative element to our understanding of the immune processes which control virus during the early critical period of infection, which can improve clinical risk stratification. The model also informs public health strategy, most notably suggesting that repeated natural exposure in sero-positive individuals may be ineffective at boosting immunity to related viral strains, and supporting the strategy of annual booster vaccinations.

## RESULTS

### A mechanistic model incorporating innate and adaptive immunity captures host/viral dynamics

We constructed a mechanistic continuous deterministic model of SARS-CoV-2 host/virus interactions based on well-accepted fundamentals of the host immune response to viral challenge. The key biological features of the model are shown in Fig 1a. Virus enters epithelial cells, replicates, is released and then infects more epithelial cells. Once infected, cells either die or stop producing virus, equivalent processes from a mathematical perspective. The parameters of epithelial cell infection have been studied in detail in a variety of in vitro models (*21, 22*), and there is a broad consensus on the kinetics. Epithelial renewal is included in the model, but death of uninfected cells is assumed to occur on a slower time scale not relevant to the model dynamics.

**Fig 1.**
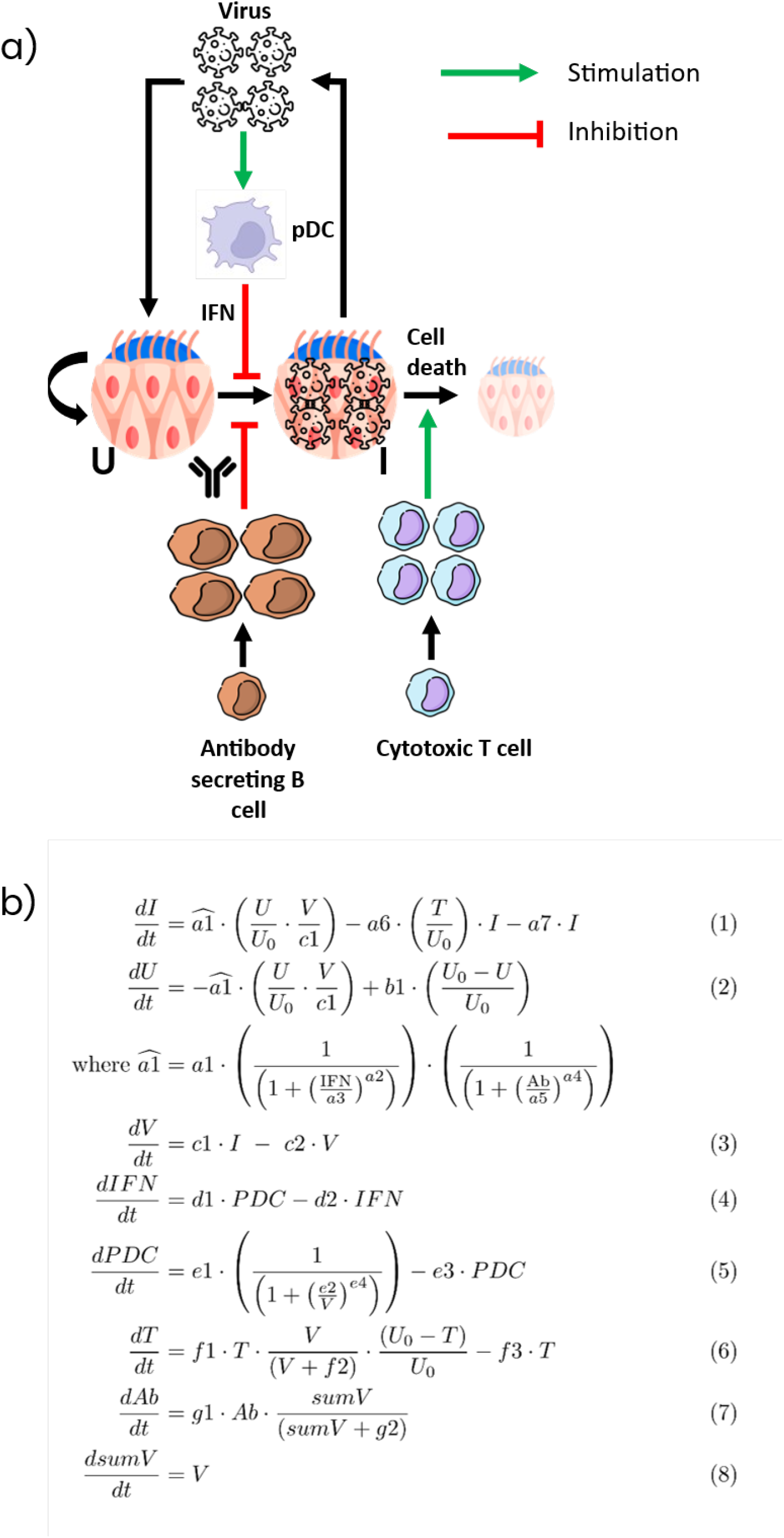
A mechanistic ordinary differential equation (ODE) model of Sars-Cov-2 infection and immune control. a) Schematic of the model. The basic infectious cycle comprises uninfected cells becoming infected by virus, and producing more virus or leaving the infectious cycle. Basal infectivity is decreased by Type I interferons (innate antiviral immunity) and antibody (virus neutralisation). The lifespan of infected cells is shortened by cytotoxic T cells. b) The 8 equations which encode the biological processes illustrated in panel a) in the model. I : infected epithelial cells; U : uninfected epithelial cells; V : free virus; Vsum : cumulative viral load. IFN : type I interferons; pDC: activated plasmacytoid dendritic cells; Ab : antibody (plasma cells producing the antibody are not modelled explicitly); T : cytotoxic T cells.

Control of viral replication is captured by three immune processes. Virus drives an innate immune response mediated by type I interferons, triggered independently from epithelial cell infection via interaction of virus and an interferon-producing cell. Interferon reduces infectivity globally across the epithelium(*5, 23, 24*). Virus also drives exponential expansion of B cells and hence plasma cells producing soluble antibody. Antibody, like interferon, reduces the infectivity rate, simulating virus neutralisation(*11, 13*). Finally, virus drives exponential cytotoxic T cell expansion(*12, 25–27*). In contrast to interferon and antibody, T cells do not affect infectivity, but increase the rate of infected cell death, simulating T cell cytotoxicity. The model is implemented as a set of ordinary differential equations (ODEs, Fig 1b) governed by a set of fixed and free parameters (Table S1). The model was parametrised by fitting to the viral load measurements from the SARS-Cov-2 challenge study as discussed in detail below.

The design of the challenge study has been described in detail previously(*15*) and is summarised in Fig 2a. In brief 34 individuals were given a fixed dose (10 TCID50, 10^6^ viral copies) of a D614G-containing pre-alpha wildtype SARS-CoV-2 (human/GBR/484861/2020; GenBank accession number OM294022). The virus was administered intra-nasally, and followed over an initial period of 28 days, by which time no viable virus was detectable. By day 90 all individuals were qPCR negative. The study design included the collection of rich and densely sampled multi-modal data for both virus and host ( Fig S1). Two basic viral replication profiles were observed (Fig 2b and Fig S2)(*15*). 18 individuals (53%) had a rapid rise in viral load within a few days of infection (PCR+), and subsequent appearance of virus specific antibody. The remaining 16 individuals (47%) had no sustained detectable virus throughout the follow up period (PCR-), no virus-specific antibody and minimal change in most other immunological measurements. In this study, we focus on the PCR+ seroconverters.

**Fig 2.**
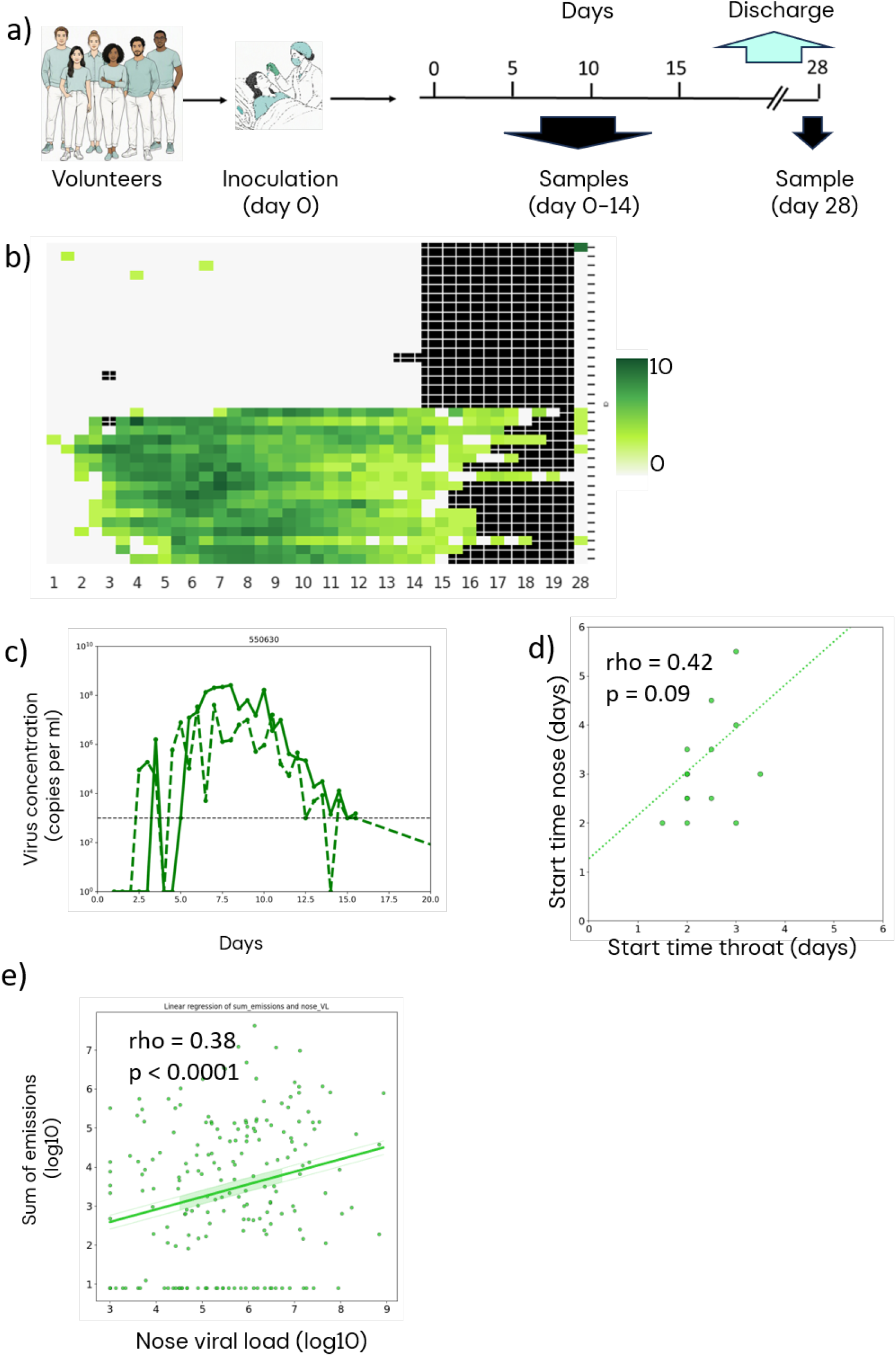
Rapid transient viraemia in a proportion of individuals receiving SARS-Cov-2 challenge. a. Schematic of the challenge study. 34 healthy volunteers received a fixed quantity of pre-alpha wildtype Sars-Cov-2 virus (10 TCID50) intra-nasally. Nasal swabs were collected twice daily for 14 days for viral load measurements, and then again at day 28 at the ned of the study. Blood samples were collected daily. b. Virus concentration measured by qPCR sampled from the nose is shown for each individual for each time point. 17 individuals showed a rapid rise in virus and then a slower fall. All individuals had undetectable virus by day 28. Black boxes are timepoints at which no sample was collected. c) A comparison of the virus profile in the nose (solid) and throat (dashed) for one representative individual. d) The time to detectable virus in throat and nose for each individual. The average time to viral detection is longer and more variable in nose, and there is no correlation between the times to detection in the two anatomical sites. e) The total amount of virus collected from the immediate vicinity of the volunteer^28^ plotted against the nasal virus concentration for each individual at each time point.

The number of viral particles was measured by qPCR from swabbing of mid-turbinates and throat every 12 hours for the first 14 days, and in some individuals for additional days sporadically until individuals were discharged. Virus was measured at days 28 (6/18 individuals still detectable in the nose) and 90 by which time virus was not detected in all individuals. A representative profile for the nose and throat is shown in Fig 2c and all individual profiles are shown in Fig S3. The trajectories follow a similar pattern; an initial very fast rise in viral titre, a period of sustained viral production over several days, and a slower return to baseline. As reported previously(*15*), on average virus replicated earlier in the throat than in the nose. The time to detectable virus varies over several days, especially in the nose, and there is no significant correlation between the time to detection in the two anatomical sites (Fig 2d, Spearman’s rho = 0.42, p = 0.09). We fit the mathematical model (Fig 1b) to the nasal viral load since the amount of virus shed (*28*), a major determinant of transmissibility, is correlated more strongly to the nasal viral load (Spearman’s rho = 0.38, p < 8*10^-8^) (Fig 2e)., than to throat viral load (0.23, p < 0.001). Since we had no information on viral replication prior to detection, we rescaled all nasal viral load data to the time of the first detectable sample prior to model fitting (see Methods).

The values for the model fixed parameters (see Table S1) were obtained from the literature as discussed in Methods. The free parameters were optimised by fitting the predicted viral load output of the model to the measured viral load measurements obtained from nasal swabs in the Sars-Cov-2 human challenge study (*15*). The free parameters of the model were determined by fitting the model’s predicted viral load to each volunteer’s nasal viral load data individually using Approximate Bayesian Inference (full details of fitting in Methods). The average (+/- standard deviation) error (i.e. difference between predicted and observed viral load) was 0.28 +/- 0.06 log10 units per measurement, in the same order of magnitude as the experimentally observed error (see Methods). The temporal predicted viral load profiles of the ten top solutions (i.e. those giving the smallest sum of squares difference from the observed virus concentrations summed over all time points) (Fig 3a) were visually very similar, illustrating convergence toward a constrained set of solutions. This is further confirmed by unsupervised clustering (Principal Component Analysis) of the parameter sets for the top 10 sets (Fig 3b). Although the parameter profiles for the different individuals differ, reflecting observed differences in viral load temporal profiles, the solutions for each individual cluster together. A plot of the distribution for the top 100 individual parameters for each individual is shown in supplementary ( Fig S4). Many of the parameters show tightly selected posterior distributions, reflecting the restraints imposed by the Bayesian posterior.

**Fig 3.**
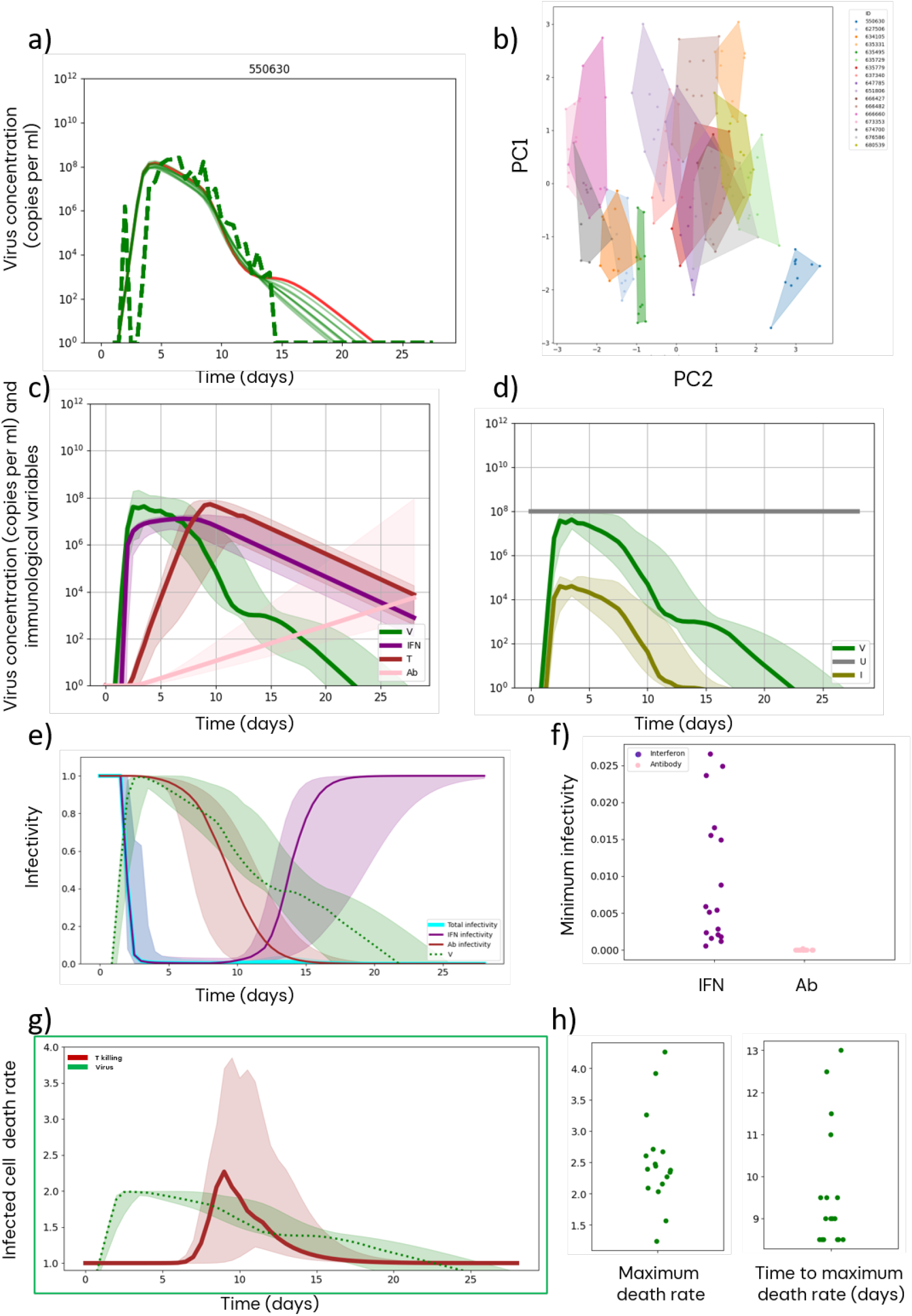
The mechanistic basis for control of viral replication in the Sars-Cov-2 model. a) The viral load profile predicted by the model with the 10 optimum (least square error) parameter sets (solid) compared to the measured viral load profile (dashed) for one representative individual. The optimum parameter set is shown in red. b) A principal component analysis (PCA) of the free model parameters for the top ten solutions. Each individual is plotted in a different colour. c) The median (solid line) and 95% confidence intervals (shaded) model predicted virus concentration (green), together with the concentrations for interferons (purple), antibody (pink) and T cells (brown). d) The median (solid line) and 95% confidence intervals (shaded) predicted virus concentration (green), together with the concentrations of infected (olive) and uninfected (grey) epithelial cells. e) The changes in total infectivity (parameter a1hat in Fig 1b, blue), and its two constitutive components which are dependent on interferon (purple) and antibody (red). Median (solid line) and 95% confidence limits (shaded). The infectivity is normalised to lie between 1 (maximum rate of infectivity) and 0 (infection completely blocked). The virus concentration profiles are shown for comparisons (green). f) The minimum value of the interferon-dependent (purple) and antibody-dependent (pink) infectivity profiles shown in panel e), plotted for each individual separately. g) The rate of infected cell death (brown) together with the virus concentrations as a function of time. h) The maximum infected cell death rate (left panel) and the time of the maximum infected cell death rate (right panel) for each individual.

The median values for the main immunological variables obtained by solving the set of equations with the optimum parameter set for all individuals are shown in Fig 3c. Individual solutions, together with measured virus profiles are shown in Fig S5.) Although differences in profiles between individuals can be observed, all share common features. The model predicts a rapid increase in interferon which parallels that of virus, followed by a short plateau and a slower fall. Antibody starts later and rises more slowly until the end of the 28 day period. T cells rise more rapidly than antibody but more slowly than interferon, and peak between days 8 and 12.

### Mechanisms which control viral replication in the model

The total uninfected cell count remains approximately constant throughout the time modelled, and only a small proportion of infectable cells actually become infected (Fig 3d). This is consistent with single cell sequencing data, which detected viral RNA in only a small proportion of epithelial cells detected in the samples even at the peak of infection(*16*) The viral restriction observed in the model is therefore not imposed by target cell limitation but to extrinsic effect of immunity. Immunity in the model controls virus by limiting infectivity (captured in the time-dependent parameter a1hat, see Fig 1b, equations 2 and 3) and by increasing the rate of T cell killing. Infectivity is the product of two components, controlled by interferon and antibody respectively. The changes in overall infectivity, and the two individual infectivity terms controlled by interferon and antibody are plotted in relation to viral concentration in Fig 3e. Infectivity (blue line) falls rapidly and subsequently remains low throughout the simulation. The component of infectivity controlled by interferon (Fig 3e, purple line) falls rapidly and effectively controls the initial viral replication but does not completely shut it off. Even though interferon reduces infectivity by 97.5-99.5 % (Fig 3f) in the model, viral replication persists because of the high natural infectivity of the virus, illustrated by the very steep initial rise in viral titres). Antibody rises more slowly, but at some point overall infectivity is limited primarily by the antibody-dependent coefficient (Fig 3e, pink line; Fig 3f). The ‘turning point’ when antibody-mediated inhibition takes over from interferon occurs around day 10-15. The rise in antibody-dependent inhibition ultimately blocks viral transmission and hence leads to viral clearance in all individuals.

In contrast to antibody and interferon which reduce infectivity, T cells in the model regulate virus by increasing the rate of viral killing (Fig 1a and Fig 1b equation 6). The rate of infected cell death is composed of a basal rate (based on in vitro data showing that infected cells stop producing virus at around one day post infection(*21*)), augmented by an additional rate due to cytotoxic T cell killing. The total infected cell death rate as a function of time and plotted alongside virus concentration is shown in Fig 3g. The increase rate of infected cell death rises as viral concentrations are already falling. While the magnitude of the effects of interferon and antibody are highly conserved in all individuals, the magnitude and kinetics of the T cell impact varies significantly between individuals (Figs 3h).

### Comparison between prediction and observed immunological activity

Having investigated the mechanisms of virus control operating in the model, a comparison between the predicted and observed immunological variables (interferon, antibody and T cells) is presented in the following section.

### Innate immunity

Type I interferon activity was measured at protein and transcriptional level. Type I interferons, as well as several interferon-inducible chemokines were detected both locally and systemically (see Fig 2 in reference(*18*)) from day 4, and peaked at day 7. Interferons are produced transiently and detection in blood and extracellular fluid is limited by sensitivity and the short half-life. However, cells exposed to interferons demonstrate a strong sustained transcriptional response (interferon responsive genes, IRGs) which has been studied in detail and can serve as a sensitive biomarker of viral infection(*19*). The temporal profile of transcription of a curated set of IRGs (list of genes is given in Table S2) in blood (Fig 4a) and in the nasal epithelium (Fig S6). A blood transcriptional IRG response was detected from day 3, peaked at days 3-4 and could be detected throughout the subsequent 14 days sampling period. The transcriptional response returned to normal levels by day 28 post infection.

**Fig 4.**
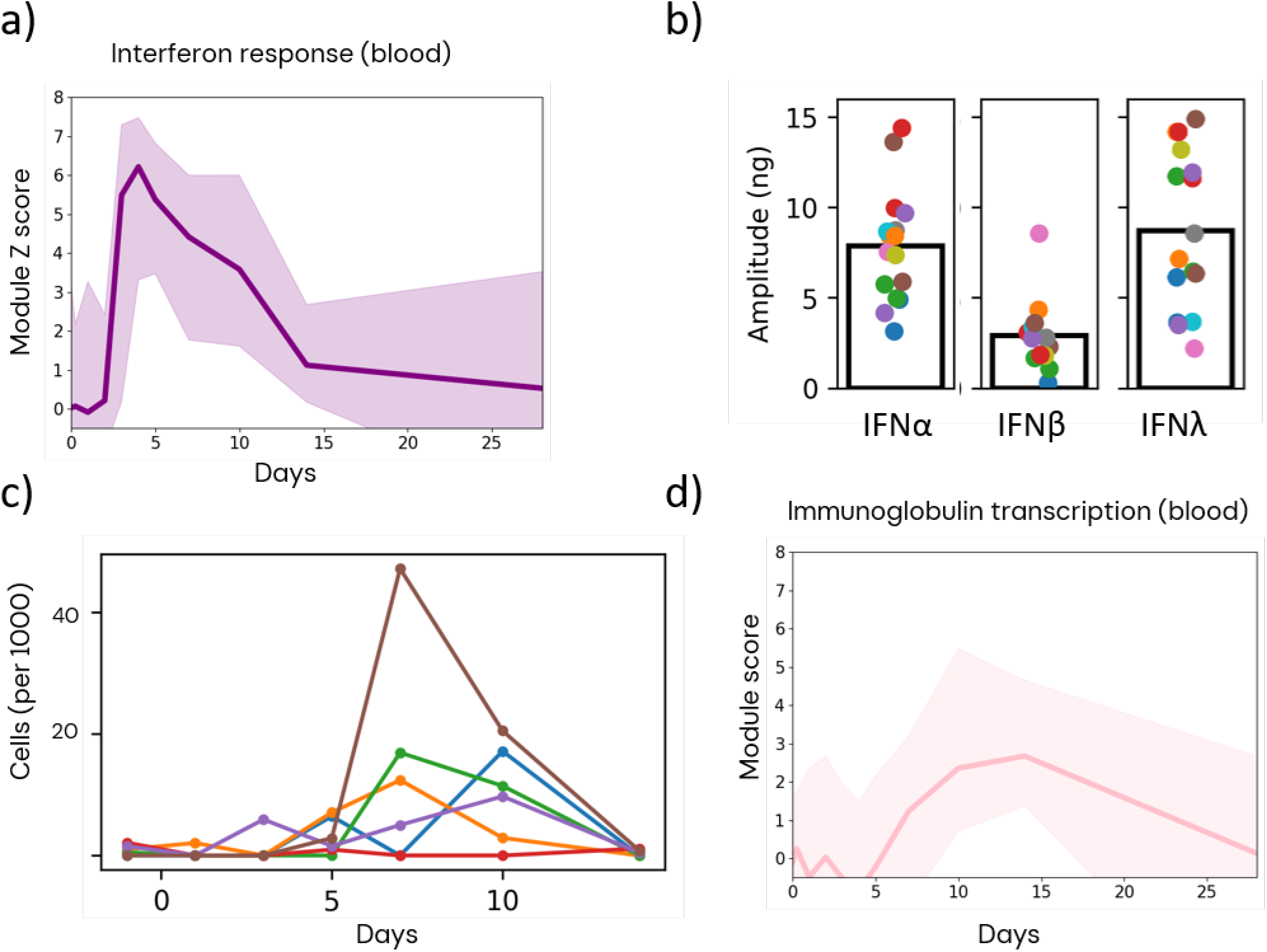
An early peak of interferon-dependent innate immunity dominates the transcriptional profile of infected individuals. a) The blood transcriptional response for an IFN responsive gene module. The data are plotted as the median transcriptional Z-score of the module (solid line) with 95% confidence intervals (shaded). The genes included in the module are detailed in Table S2. b) Interferon α, β and λ (IL29) protein levels in blood, measured by ELISA. Each dot represents one volunteer. c) The proportion of pDC observed in the nasal epithelium, as measured by single cell transcriptional profiling as described in detail elsewhere^16^. Each colour is one individual. d) The blood transcriptional response of an immunoglobulin transcription gene module. The data are plotted as the median transcriptional Z-score of the module (solid line) with 95% confidence intervals.

The Type I interferons detected were predominantly IFNα and IFNλ (IL29), while IFNβ levels are lower or undetectable (Fig 4b). Interestingly, this pattern of type I interferon response is characteristic of plasmacytoid DC (*29*), while epithelial cells produce a robust IFNβ response. The kinetics of the interferon response are compatible with a small but consistent increase in pDC signal in the single cell nasal data detected between days 5-7 (Fig 4c). It is also consistent with the absence of a strong transcriptional signal for any of the type I interferons, since pDC constitute only a small proportion of cells, but individual cells can produce enormous quantities of protein. Finally interferon production by pDC, which are not themselves productively infected, also reconciles the apparent paradox that we observe a strong and rapid interferon response, despite potent SARS-CoV-2 mechanisms for inhibition of interferon production (*23, 24, 30*).

Taken together, the observed protein and transcriptional pattern of interferon activity is consistent with the structure and output of the mathematical model, reflecting a strong rapid innate immune response peaking more or less at the same time as virus, and decreasing slowly as virus levels fall. The strength of viral inhibition in the presence of interferon alpha observed in vitro, and the observation that interferon restriction can be partially overcome by high virus concentrations is also consistent with the inhibitory activity predicted by the model (*21, 23, 30*).

### Antibody mediated immunity

The antibody response in the challenge study has been reported previously(*15, 18*). Specific serum antibody can first be detected at day 10 (IgM and IgG are more or less contemporaneous) and continues to rise until the end of the study. Neutralizing antibody is detected at day 14. Transcriptional profiling revealed a transient increase in immunoglobulin transcription detected at day 6, and remaining above basal level until day 14 (Fig 4d). We hypothesise this reflects the transient appearance of B cell plasmablasts in the blood, as previously reported using flow cytometry (*18*). These kinetics are consistent with the model predictions, which suggest that antibody neutralisation becomes the major factor regulating infectivity after day 14 post-infection (Fig 3e).

### T cell immunity

Previous studies using flow cytometry and interferon-release assays documented a SARS-Cov_2 specific CD8 T cell response on days 10-14 following infection(*18*). Consistent with these previous observations, we observed a T cell activation and proliferative transcriptional signal peaking at the same time in blood (Fig 5a). In the nasal epithelium transcriptome, a local increase in activated T cells was also observed around days 7-10, but there was no evidence of a proliferative signal (Fig 5b), suggesting that the recruitment of activated T cells to the epithelium is driven predominantly by migration rather than proliferation in situ.

**Fig 5.**
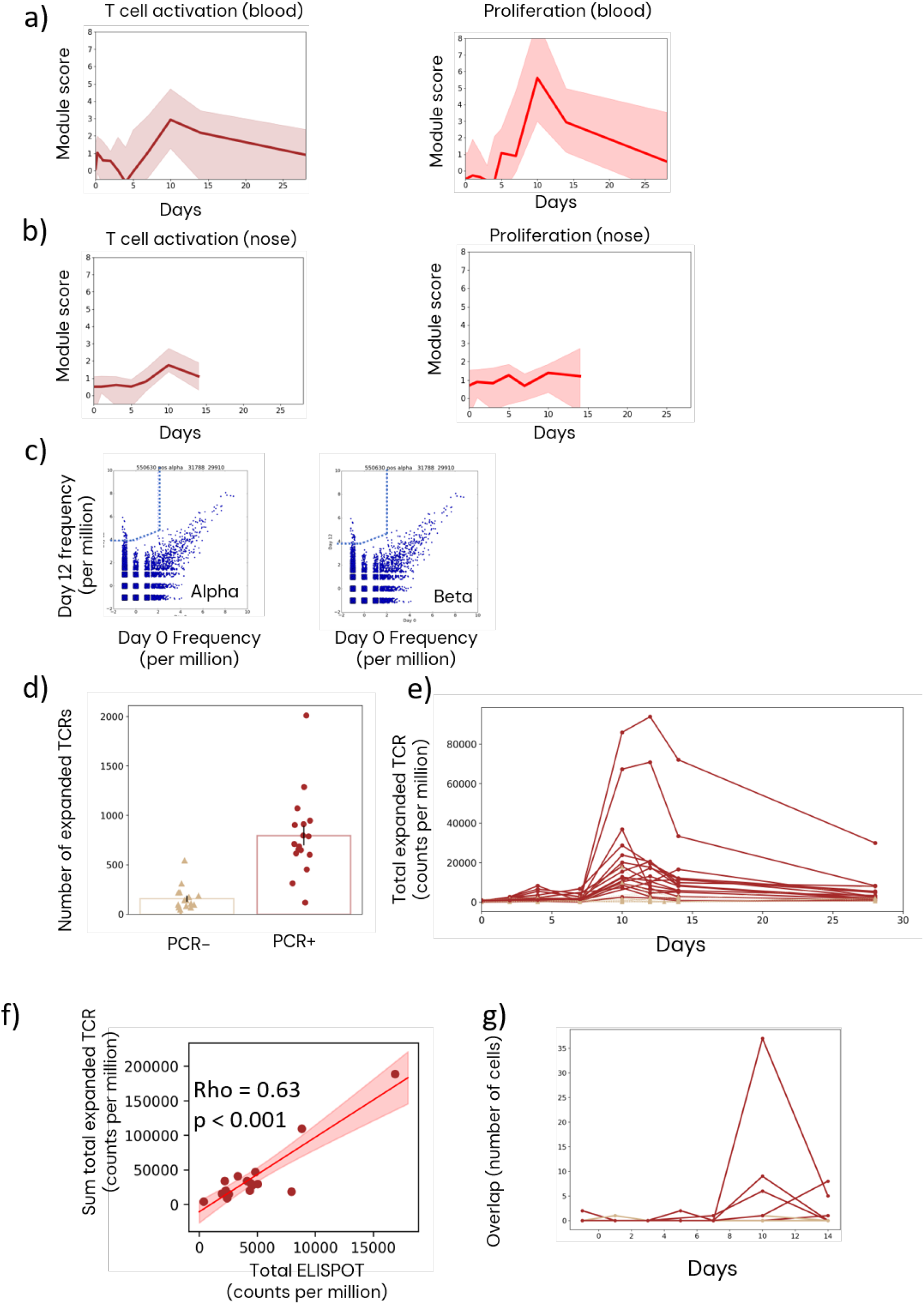
Clonal expansion of T cell receptors following infection with Sars-Cov-2. a) The blood transcriptional response of the T cell activation module (left) and proliferation module (right). The data are plotted as the median module transcriptional Z-score (solid line) with 95% confidence intervals (shaded). The genes included in the module are detailed in Methods. b) As for a, but for the nasal epithelial transcriptome. c) The frequency of TCRα and TCRβ at day 12 (y axis) versus the frequency at day 0 (x axis) for all TCRs from the TCR repertoire of one individual. Each dot represents a unique TCR sequence. The TCRs in the upper left region (ULR) are significantly expanded at the later time point (see Methods for statistical details). d) The number of expanded TCRα chain sequences at any time post day 4 in volunteers who do or do not develop a productive infection (cf Fig 2b). e) The sum of the TCRα chain clonal frequencies for each individual as a function of time in volunteers who do (brown) or do not (tan) develop a productive infection. f) The total frequency of TCRα is correlated to the SARS-Cov-2 specific IFN-γ ELISpot count. The TCR frequencies and the ELIspot data were summed over all days for which data was available. The details of the ELISpot assays are given in ^17^ g) The number of expanded TCRα chains observed in both blood and nasal epithelium. The TCR sequences from the nasal epithelium were obtained by single cell transcriptomics as described in detail in ^16^.

We sequenced the TCRalpha and TCRbeta repertoire of whole blood samples at days 0, 2, 4, 7, 10, 12, 14 and 28 of the challenge. We then compared the clonal frequency of each TCR between days zero (samples taken pre-infection) and all other days, and identified TCRs which increased significantly between the two time points (Fig 5c). We refer to these as expanded TCRs. The number of expanding TCRs responding in each individual in productively infected and non-infected individuals is shown in Fig 5d and Fig S7a. The sum of the clone count for all expanding TCRs at each time point for each individual provides a surrogate for the strength of the T cell response (Fig 5e and Fig S7b). In productively infected individuals, the T cell response peaks at day 10 – 12, consistent with the transcriptional signals in Fig 5a. T cell expansion then falls more gradually, but remains above the pre-challenge values at day 28. The total frequency of expanding T cells also correlates with the gamma interferon response measured by ELIspot (Spearman correlation = 0.5, p =0.03)(Fig 5f). We also compared the expanding TCRs observed in blood with single cell transcriptomic data obtained from T cells in the nasal epithelium available for 5 individuals (*16*). The time course of the matched TCRs in the nasal epithelium reflected the abundances of the T cell expansions seen in blood, and peaked at day 10 (Fig 5g and Fig S8). On day 10 we detected exact matches between TCR sequences of intra-epithelial T cells and expanding TCRs for 50/1847 cells (2.7 %) TCR alpha and 67/2328 cells (2.8%) TCR beta sequences.

In summary, both global transcriptomics and TCRrep sequencing detects a population of rapidly expanding T cells in blood 10-14 days post infection, consistent with the immunological outputs of the model. The data obtained from the site of infection suggest that the expanded T cells migrate rapidly, but transiently into the nasal epithelium.

### Model perturbation predicts the impact of immunodeficiency

A major strength of a mechanistic model is it can be used to explore predictions about the host/virus interaction under different scenarios. We first perturbed the model by decreasing the amount of interferon secreted, the production of antibody and the amount of T cell killing (Fig 6). Decreased interferon production (Fig 6a, b) is accompanied by a rise in total and maximum virus production which reaches more than 1000 fold (Fig 6c). Decreased interferon also increases the time to viral clearance (Fig 6d), but as adaptive immunity develops, virus replication reaches undetectable concentrations in all individuals. These predictions are supported by the observations of increased viral loads and higher rates of hospitalization, but eventual viral control in individuals with genetic defects in the interferon-response pathway or in the presence of anti-interferon auto-antibodies(*31–34*).

**Fig 6.**
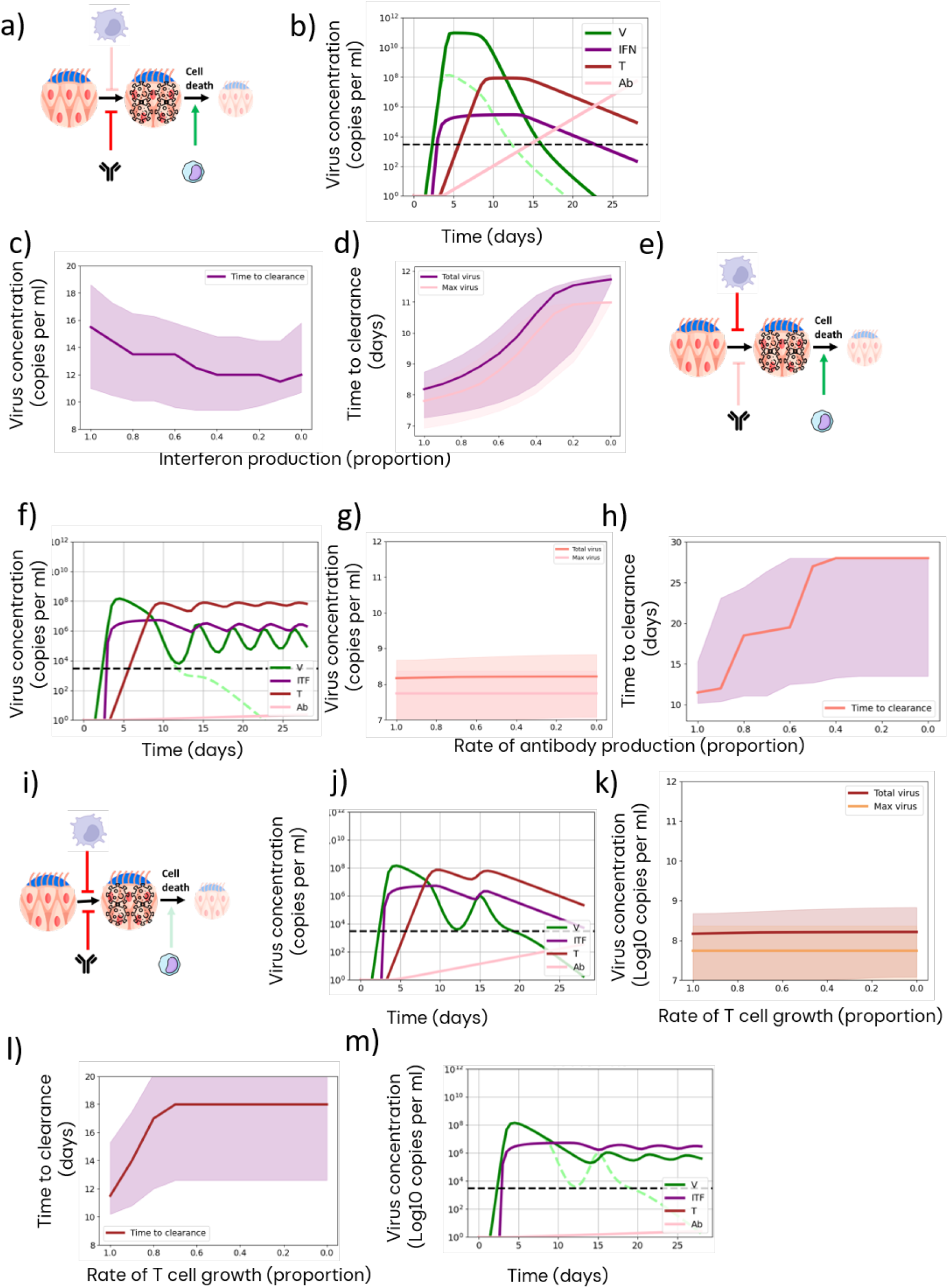
The mathematical model predicts the impact of immunodeficiency on Sars-Cov-2 magnitude and dynamics. a) A representative model schematic of type I interferon deficiency in the model. b) An example of viral (solid line, green), interferon (purple), cytotoxic T cell (brown) and antibody (pink) kinetics when interferon production (parameter d1 in Fig 1b) is reduced by 90%. Virus kinetics in the unperturbed model is shown by the dashed green line. The detection level for virus is shown by the dashed black line. c) The impact of decreasing interferon production on maximum (salmon) and total (area under the curve, pink) virus load. The plot shows median (solid lines) and 95% confidence intervals (shaded). d) The impact of decreasing interferon production on the time at which virus is cleared, which is set as the time the concentration falls to the detection limit of 3000 copies per ml. The plot shows median (solid lines) and 95% confidence intervals (shaded). e) A representative model schematic of antibody deficiency in the model. f) An example of virus (solid line, green), interferon (purple), cytotoxic T cell (brown) and antibody (pink) kinetics when antibody production (parameter g1 in equation 7, Fig 1b) is reduced by 90%. Virus kinetics in the unperturbed model is shown by the dashed green line. g) The impact of decreasing antibody production on maximum (salmon) and total (area under the curve, pink) virus load. The plot shows median (solid lines) and 95% confidence intervals (shaded). h) The impact of decreasing antibody production on the time at which virus is cleared, which is set as the time the concentration falls to the detection limit of 3000 copies per ml. The plot shows median (solid lines) and 95% confidence intervals (shaded). i) A schematic of cytotoxic T cell deficiency in the model. j) A representative model example of virus (solid line, green), interferon (purple), cytotoxic T cell (brown) and antibody (pink) kinetics when T cell growth (parameter f1 in equation 6, Fig 1b) is reduced by 90%. Virus kinetics in the unperturbed model is shown by the dashed green line. k) The impact of decreasing T cell growth on maximum (salmon) and total (area under the curve, pink) virus load. The plot shows median (solid lines) and 95% confidence intervals (shaded). l) The impact of decreasing cytotoxic T cell growth on the time at which virus is cleared, which is set as the time the concentration falls to the detection limit of 3000 copies per ml. The plot shows median (solid lines) and 95% confidence intervals (shaded). m) The impact of a 90% decrease in T cell growth in the presence a 50% decrease in antibody production.

In contrast, inhibition of antibody (Fig 6e, f) had minimal impact on the initial virus peak, and on total maximum virus concentration (Fig 6g). However, a decrease in antibody had a dramatic impact on viral clearance, ultimately leading to sustained viral production throughout the 28 day period (Fig 6h). Despite this increased persistence the total viral load summed over the 28 days was not increased much, because the bulk of the virus production occurred during the early peak. The prolonged and persistent levels of viral production observed in individuals with antibody deficiencies (*32, 35, 36*) are consistent with these predictions.

Decrease of T cell killing in the model (Fig 6i, j) showed a less severe phenotype, with minimal effects on the initial peak, or on total or maximal viral concentration (Fig 6k), but a moderate and variable increase in time to clearance (Fig 6l). These predictions suggest cytotoxic T cells are secondary to antibody in protecting against acute infection, but may play a role in shortening the viraemic peak(*18*). In order to probe the role of T cells further, and since impaired antibody responses are often associated with impaired T cells responses, we investigated the impact of decreasing T cell responses under conditions of low antibody production. Under conditions of low antibody levels, the role of T cells on viral clearance is exaggerated and T cells become essential for controlling chronic viremia (cf Fig 6 j and m).

### Model perturbation predicts the impact of prior immunity

We explored our model further by introducing pre-existing immunity. In the presence of increased pre-existing antibody (Fig 7a, b), virus replication was inhibited (Fig 7b) and above an antibody threshold abolished altogether. The level of antibody required to block viral replication before exposure was less than the amount required for viral clearance in the unperturbed model (cf Fig 6b), reflecting the low concentration of virus early post-challenge. One striking prediction of the model is that the boosting effect of viral re-exposure becomes smaller and slower (Figs 7c, d), since innate and adaptive immunity are driven by the increase in virus concentration. We compared these predictions to the measurements obtained during a second challenge study(*20*), where sero-positive individuals were challenged with the delta strain of virus. In agreement with the model, viral replication was restricted in the presence of pre-existing antibody, and only observed when volunteers were pre-selected for low titres of antibody. Even in this restricted cohort, only 33% of individuals became infected. In individuals in whom viral replication was absent, there was minimal boosting of either antibody or T cell activity. In those individuals who did become infected, viral titres were much lower than in the seronegative challenge study (Fig 7e), modest antibody boosting and a much reduced T cell response were observed. We measured the blood transcriptional response following challenge in this second study (Figs 7f-h). Both the interferon and the immunoglobulin transcriptional responses were substantially reduced compared to the first challenge (Figs 7f,g) and the T cell activation transcriptional module was undetectable (Fig 7h). In contrast to the lack of boosting from a challenge dose of virus, the model predicted strong immunity if initially stimulated with a larger bolus of virus, such as might occur during vaccination (Fig 7i). The model therefore predicts that vaccination is required to optimally boost immunity to similar viruses in sero-positive individuals, but natural infection may not be effective. Infection with very antigenically different virus strains (e.g. omicron strain), which are not blocked by the pre-existing antibody, may still be effective at boosting immunity.

**Fig 7.**
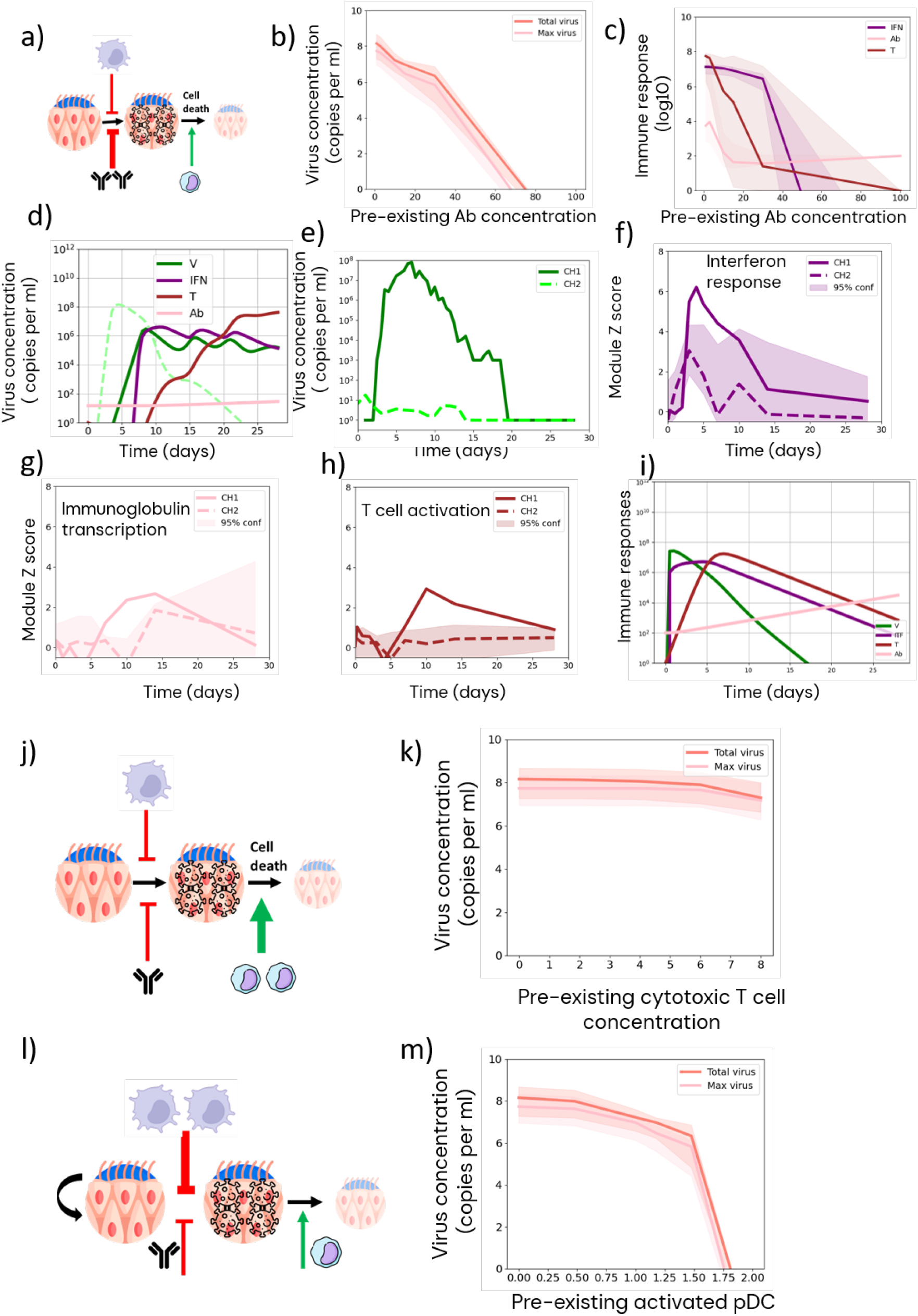
The mathematical model predicts the impact of pre-existing immunity on Sars-Cov-2 magnitude and dynamics. a) A model schematic of pre-existing antibody in the model. b) The impact of increasing levels of pre-existing antibody on maximum (salmon) and total (area under the curve, pink) virus load. The plot shows median (solid lines) and 95% confidence intervals (shaded). c) The impact of increasing levels of pre-existing antibody on the maximum interferon (purple), antibody (pink) and cytotoxic T cell levels. d) An illustrative example of the impact of pre-existing antibody (50 units) on virus (green), interferon (purple), cytotoxic T cell (brown) and antibody (pink) kinetics. Virus kinetics in the unperturbed model is shown by the dashed green line. e) The nasal viral load observed in the second seronegative delta variant challenge (dashed line) compared to the first seronegative challenge (solid line). f) The interferon-dependent transcriptional signature in the second seronegative delta variant challenge (dashed line) compared to the first seronegative challenge (solid line). g) The immunoglobulin transcriptional signature in the second seronegative delta variant challenge (dashed line) compared to the first seronegative challenge (solid line). h) The activated T cell transcriptional response in the second seronegative delta variant challenge (dashed line) compared to the first seronegative challenge (solid line). The plot shows the median transcriptional Z-score of each module (solid line) with 95% confidence intervals (shaded). The genes included in the modules are detailed in Methods. i) The interferon (purple), antibody (pink) and cytotoxic T cell response following an initial exposure to a bolus of virus (green). j) A model schematic of pre-existing cytotoxic T cells in the model (left), and the predicted impact of increasing levels of pre-existing levels of cytotoxic T cells on maximum (salmon) and total (area under the curve, pink) virus load. The plot (right) shows median (solid lines) and 95% confidence intervals (shaded). k) A model schematic of pre-existing innate immunity in the model (left) and the predicted impact of increasing numbers of pre-activated pDC on maximum (salmon) and total (area under the curve, pink) virus load. The plot (right) shows median (solid lines) and 95% confidence intervals (shaded).

In contrast to the strong inhibitory effects of pre-existing antibody, it was difficult to prevent infection, or even substantially reduce viral loads in the presence of pre-existing T cell activity but without pre-existing antibody, even at high levels (Fig 7j). A pre-existing innate response (Fig 7k), on the other hand, was effective at decreasing the viral load, as recently reported for several respiratory infections in mouse *in vivo* studies (*37*)

## DISCUSSION

The SARS-CoV-2 pandemic stimulated an enormous number of studies describing the host immune response to viral infection. Nevertheless, an integrative quantitative model of the human response to SARS-CoV-2 is still lacking. In this study we encode prior knowledge about the fundamental features of innate and adaptive immunity into a mechanistic deterministic model which predicts not only the viral dynamics with which the model was parametrised, but also experimental measurements of innate and adaptive immunity obtained in a SARS-Cov-2 human challenge study. The focus on explicit modelling of multiple immune components, and the link to densely sampled experimental data from the very earliest phases of infection distinguish the study from several previous mathematical models of SARS-Cov-2 in host dynamics (see review in (*10*)). Model perturbation is applied to predict the impact of defects in innate or adaptive immunity, and the impact of pre-existing immunity on viral dynamics and immune boosting. The model highlights the role of incomplete suppression of viral replication by early innate immunity in the large sustained peak of virus following infection with SARS-Cov-2, and the critical switch from innate to adaptive immunity which drives viral clearance in most healthy individuals.

Mathematical modelling allows us to test our intuition about the fundamental mechanisms which protect the host from uncontrolled virus replication. In the context of SARS-CoV-2 infection, we isolate three principal strands of the host response, interferon-mediated innate immunity, T cell cytotoxicity and antibody-mediated neutralisation. After fixing parameters which can be estimated from the literature, we constrain the remaining model parameters to fit the viral load dynamics from the SARS-Cov-2 challenge study, discarding all the immunological data from this study. We opted to fit each individual separately, to exploit the variability we observe in viral dynamics and to begin to explore the extent of inter-individual heterogeneity. Despite the number of parameters, the models converge towards a very similar common solution in all the individuals. The parameters show weak inter parameter correlations, suggesting that each parameter is contributing non-redundantly to the model. The optimum parameters cluster into different areas of a two-dimensional projection, suggesting that the different viral trajectories represent slightly different solutions for the immune response in different people.

Despite the inter-individual variability, a clear shared pattern of host/virus interaction emerges in all the individuals. A strong Innate immune response emerges rapidly and almost contemporaneously with virus, in both model and experimental data. However, the model suggests that, because of the very high rate of virus reproduction, innate immunity reduces but does not completely block viral replication/transmission, which results in a high viral load maintained over several days. It is only as antibody and cytotoxic T cells appear, around days 10-12, that viral load falls rapidly becoming undetectable from around day 14 in most individuals. The model suggests that antibody neutralisation, rather than T cell cytotoxicity plays the dominant role in this process. The mechanistic reason for this, in the model, is that antibody neutralisation decreases infectivity directly, while the T cells can only increase the rate of epithelial cell death after they are infected. Furthermore, since T cells and their targets need to interact directly to result in target cell death, the cytotoxic effect is dependent on the product between concentration of infected cells and concentration of T cells and is therefore only effective at high T cell numbers. Nevertheless, the model predicts that T cell activity does shorten the length of time until virus becomes undetectable in some individuals, consistent with the observed correlation between CD8 T cell activity and viral clearance(*18*).

A non-redundant dominant role for antibody in viral clearance is also seen when perturbing the model by decreasing the rate of antibody production, and is consistent with several reports of long term persistent viremia in infected individuals with antibody deficiency (*38, 39*). However, the major peak of viremia is observed early in infection, before antibody or T cells become active. The magnitude of this initial viral peak is believed to be the major determinant of progression to long term morbidity and mortality, which can occur long after virus load becomes low or undetectable. The magnitude and persistence of this early viral peak is determined in the model by the effectiveness of innate immunity, and even partial abrogation of interferon production results in an increase both in the maximum and total levels of viremia observed. These model predictions are consistent with the well-established link between a decreased type I interferon response, and increased early viral load and increased progression to serious disease(*31–34, 40*). The model also suggests that the effectiveness of the interferon response may be a key factor determining transmissibility of SARS-Cov-2, since the total viral load in the nose is correlated to viral emissions(*28*). However, we have not yet formally integrated our in-host model with inter-host transmission models.

The model can also be used to generate testable hypotheses about the role of individual components of pre-existing immune responses, generated either by prior exposure or by vaccination in preventing, or diminishing infection. In this setting, as in immune naïve individuals, the model points to a dominant role for neutralising antibody. The correlation between neutralising antibody levels and protection against SARS-Cov-2 infection is now well-established. For example, in the second SARS-Cov-2 (delta variant) challenge study, carried out in seropositive individuals, detectable virus was observed only in individuals preselected for lower levels of neutralising antibody prior to exposure, and even in this restricted group replication was observed in only 33% of the volunteers(*20*). However, there is increased interest in vaccines focused on T cell responses(*41*), and a recent study suggested that priming of innate immunity alone may provide broad protection against multiple virus types(*37*). The model predicts that T cell cytotoxic immunity would only provide effective protection if immunity could be maintained at high levels over long periods of time. Indeed ‘breakthrough’ immunity could even give rise to delayed but enhanced viral replication, since the delay in peak viraemia would also delay innate and antibody-dependent immune responses. The model suggest that strategies aimed at inducing cytotoxic T cells should therefore be explored with caution, but supports further exploration of strategies for boosting innate immunity.

The model also highlights the two-way interaction between virus and host. In the discussion above, we focus on the role of immunity in controlling viral replication. However, virus has a key role in stimulating adaptive immunity, which is highlighted under conditions where pre-existing immunity limits the initial virus replication. The key message here is that an existing effective immune response prior to virus exposure will not only prevent viral replication but will prevent or significantly attenuate and delay immune boosting since the virus antigens do not reach concentrations high enough to activate the immune response. The model therefore suggests that natural ongoing exposure in protected individuals will not necessarily replace the role of repeated vaccination in boosting immunity. These predictions are supported by the greatly reduced and delayed immune responses observed in a second SARS-Cov-2 delta variant) challenge study, carried out in sero-positive individuals. This two-way interaction also highlights the challenges of imputing causality to observed correlative relationships between immune responses and viral dynamics.

The study has a number of limitations. On the experimental side, the challenge study is restricted to a small number of healthy volunteers. The data is therefore focused on a protective response and does not address the later immune events which accompany more serious morbidity or mortality. On the computational side, the modelling is restricted to ODE-based deterministic and continuous models. Future studies could incorporate stochastic elements, especially to capture the early events post infection where very few cells are infected. The model does not investigate spatial segregation, which can be incorporated using partial ODEs. The model also simplifies the immunology in favour of interpretability and computational robustness. Future refinements could include the role of macrophages, NK cells and other cell types implicated in SARS-CoV-2-specific immunity(*42*).

In conclusion, we present an integrative analysis of SARS-CoV-2 immunity, in the context of a human challenge study of a cohort of young healthy virus-naïve individuals. In those individuals who develop productive infections the results highlight that despite significant inter-individual variation in many components of innate and adaptive immunity, a shared regulated sequence of immunological responses lead to control and clearance of virus in all the individuals tested. The overall sequence of immunological responses can be captured by a simplified mechanistic and deterministic model which combines interferon-driven innate immunity, cytotoxic T cells and antibody. Our interpretation of the model has three important translational impacts. First, it consolidates our focus on innate immune and antibody deficiencies as the critical risk factors for severe disease and chronic infection. Second, it suggests vaccination strategies focussing on cytotoxic T cell boosting are likely to have limited success in 14the absence of neutralising antibodies. Third, the model informs public health policy by highlighting the modest immune boosting achieved by low dose natural exposures, necessitating repeated vaccination to maintain high levels of protective immunity. These findings may be expected to be generalisable to other acute respiratory viruses.

## MATERIALS AND METHODS

### Modeling

#### Boundary conditions for ODEs

The number of starting T cells, B cells (antibody producing cells) and pDC were arbitrarily set to 1 in the default model. The number of infected cells was set to zero and the number of initial uninfected cells set to 10^8^, estimated as the number of epithelial cells in the nasal epithelium, calculated by dividing the surface area of the epithelium(*43*) by the surface area of the epithelial cells(*44*). Similar numbers have been used to model infection of the nasal epithelium previously(*45, 46*). The changes in viral concentration prior to detection are unknown. We therefore set the initial viral concentration at day 0 to 10^-5^ which is the approximate estimated viral concentration at day zero, assuming a single initial exponential phase as in the model. The sensitivity of the model to varying the initial viral concentration is shown in Fig S9a.

The set of parameters for the model are summarised in Table S1. The fixed parameters were determined as follows.

1. a4. Antibody neutralisation Hill coefficient = 2; many antibodies to SARS-Cov-2 demonstrate positive co-operativity with Hill coefficients in the range of 1.5 – 4. Changing the Hill coefficient between 2 and 4 had only a small impact on model output (Fig S9b).
2. a5 Antibody neutralisation IC50 = 10; the units of antibody concentration are arbitrary, and the IC50 and the rate of antibody production are not independently determined. Fixing one of these, and optimising the other allows unambiguous model selection.
3. a7 The rate of infected cell death = 1 day^-1^. This parameter captures the transition from a cell producing virus to a cell no longer producing virus; the model does not distinguish whether this occurs because the cell dies or simply ceases producing virus. We estimate this value from in vitro studies on virus production(*21*). This estimate is close to the value obtained from an independent modelling study where this parameter was free, and inferred from the data fitting(*47*). The sensitivity of the model to varying this parameter is shown in Fig. S9c
4. b1 The rate constant of replenishment of epithelial cells = 10^8^ d^-1^. Density dependent epithelial replacement does not play any role under in the default model, because only a very small proportion (<1%) of the epithelial cells become infected. However, under conditions of chronic infection, when immunity is impaired, the term prevents epithelial cells from being completely depleted. The rate constant is set such that the epithelial cells replace at 10^6^ day when the number of uninfected cells drops by 1%.
5. c1 The rate of infectious virus production = 10000 infectious virions cell^-1^d^-1’^ This was estimated from in vitro studies of virus production in cultured epithelial cells(*21*). The sensitivity of the model to varying this parameter is shown in Fig. S9d
6. c2 Rate of virus clearance = 10 day^-1^. This is similar to the rates of virus clearance estimated both for SARS-Cov-2 and Influenza (*46, 47*). It is consistent with the very fast rate of clearance at the nasal mucosal surface(*48*). The model output is insensitive to the precise value of the rate, provided it is significantly greater than the death of infected cells.
7. d1 Rate of interferon production = 10 cell^-1^day^-1^; the units of interferon production are arbitrary, and the production rate and IC50 of interferon inhibition are not independent. Fixing one of these arbitrarily (thus determining the unit scale), and optimising the other allows unambiguous model selection.
8. d2 Rate of interferon clearance = 10 day^-1^; set at the same value as viral clearance, assuming the rate of clearance is set by the rate of mucosal flow over the epithelial surface(*48*). Interferon clearance may be even faster, but increasing the rate does not impact the model output.
9. e3 Rate of pDC death = 0.5 day^-1^. Based on in vivo estimates of pDC turnover. (*49, 50*) The sensitivity of the model to varying this parameter is shown in Fig. S9e
10. f3 Rate of cytotoxic T cell death = 0.5 day^-1^. Based on in vivo estimates of the rapid contraction phase of cytotoxic T cell responses to primary SARS-Cov-2 infection. The model does not incorporate emergence of memory T cell responses. The sensitivity of the model to varying this parameter is shown in Fig. S9f.

The remaining parameters were selected by fitting the model to the observed nasal viral load using Approximate Bayesian Inference. The model was fitted to each individual separately, to obtain estimates of parameter variation. Since there was no information on the events occurring prior to virus becoming detectable, and since it was observed that the time to viral detection was variable (Fig 2d) suggesting some stochastic process at the earliest stages of the infection when the number of infected cells was very small, the viral load data was shifted along the time axis so that time to the first detectable virus was 2 days (the minimum delay observed). The set of equations (Fig 1b) was solved 10^6^ times, each instance randomly and uniformly selecting a new set of parameters from within the range shown in Table S1. For each solution, the root mean squared error between observed and predicted virus concentration (Log10 viral load measurements) was calculated (two measurements per day were available). The solutions were ranked in increasing order of error, and the thousand solutions with the lowest error were selected. The range of parameter values allowed were reset to the 90^th^ percentiles of the values observed in these thousand solutions, and then the equations were solved a further 10^6^ times. The solutions were again ranked in ascending order of root squared mean error between predicted and observed values, and the solutions with lowest error were selected for further study. The average error observed falls by 90% of that observed with no Bayesian constraints after the first round of Bayesian selection, but only by 1-2% more after the second round of Bayesian selection (Fig S10). As shown in Fig S4 several parameters were tightly selected in the posterior distribution, indicating a strong dependence of the model on the parameter. There was only rather weak correlation between the parameters, suggesting the impact of the variable parameters on the model were independent and non-redundant (Fig S11).

#### The challenge models

The first Sars-Cov-2 Challenge study and sample collection from previously seronegative individuals has been described in detail previously(*15, 16, 18*). The second challenge study is described here (*20*)

#### Viral load measurements

The number of viral particles was measured by qPCR, sampling by swabbing the nasal (every 12 hours) and throat (daily) epithelium for the first 14 days, and in some individuals for additional days sporadically until virus was undetectable. Virus was also measured at days 28 and 90 but was undetectable in all individuals at these time points.

As our goal was to generate a model which quantitatively fitted virus dynamics, it was important to estimate the error in the viral load measurements, which could arise from qPCR measurement or sampling. Duplicate qPCR measurements, carried out in different laboratories showed a high degree of reproducibility, with a median difference of 0.15 log10 units (Fig S12). We did not have access to duplicate sampling, so we estimated an upper bound to sampling error, by measuring differences between successive 12 hour time points (Fig S13). The median absolute difference between viral load at successive time points was 0.7 Log10 units. We also estimated the empirical PCR sensitivity threshold by calculating the distribution of minimum PCR values. The median was 3 log10 units.

#### Transcriptomics

Whole blood RNA was collected and processed as described in (*19*). Processed RNAseq data is available at ArrayExpress for the SARS-CoV-2 challenge study (accession number: E-MTAB-12993, https://www.ebi.ac.uk/biostudies/arrayexpress/studies/E-MTAB-12993). To comply with data privacy restrictions, raw sequencing data is available under managed access through the European Genome-Phenome Archive (https://ega-archive.org), under the following accession numbers: EGAD50000000942 for SARS-CoV-2 challenge study (https://egaarchive.org/datasets/EGAD50000000942). The genes which make up the individual modules are given in Table S2. For each module, module scores were calculated as arithmetic mean expression of the constituent genes, using log2 transcript-per-million values.

- *TCR sequencing and analysis.* The α and β chains of the TCR repertoire were sequenced from the same RNA sample as used for the global transcrptomics. Samples from days 0, 2, 4, 7, 10, 12, 13, 14 and 28 from all individuals who were PCR+ for at least three consecutive time points were processed for TCRseq using an in house pipeline. The pipeline introduces unique molecular identifiers attached to individual cDNA molecules using single-stranded DNA ligation The UMI allows correction for sequencing error PCR bias, and provides a quantitative and reproducible method of library preparation(*51, 52*). The raw sequences were parsed using the in-house software suite Decombinator (*53, 54*). The software is open source and can be found at https://github.com/innate2adaptive/decombinator.

Expanded TCRs were defined as TCRs which changed significantly between any two time points. The significance boundaries (shown as blue dotted lines in Figure 4a) were defined as the maximum TCR abundance which might be observed at time 2, given its abundance at time 1, given Poisson distribution of counts with p < 0.0001, to give a False Discovery Rate of <1 in 1000. TCR abundances are normalised for total number of TCRs sequenced in each sample, and expressed as counts/million.

## Data Availability

All data sets and files used in this study are deposited at 10.5281/zenodo.22832153. All code for analysis and generating individual figure panels are available on GitHub https://github.com/innate2adaptive/Modelling-SARS-COV-2-challenge/ The analysis was carried out using Python v3.10.12. Any additional information required to reanalyse the data reported in this paper is available from the lead contact upon request.

## List of Supplementary Materials

**Fig S1.**
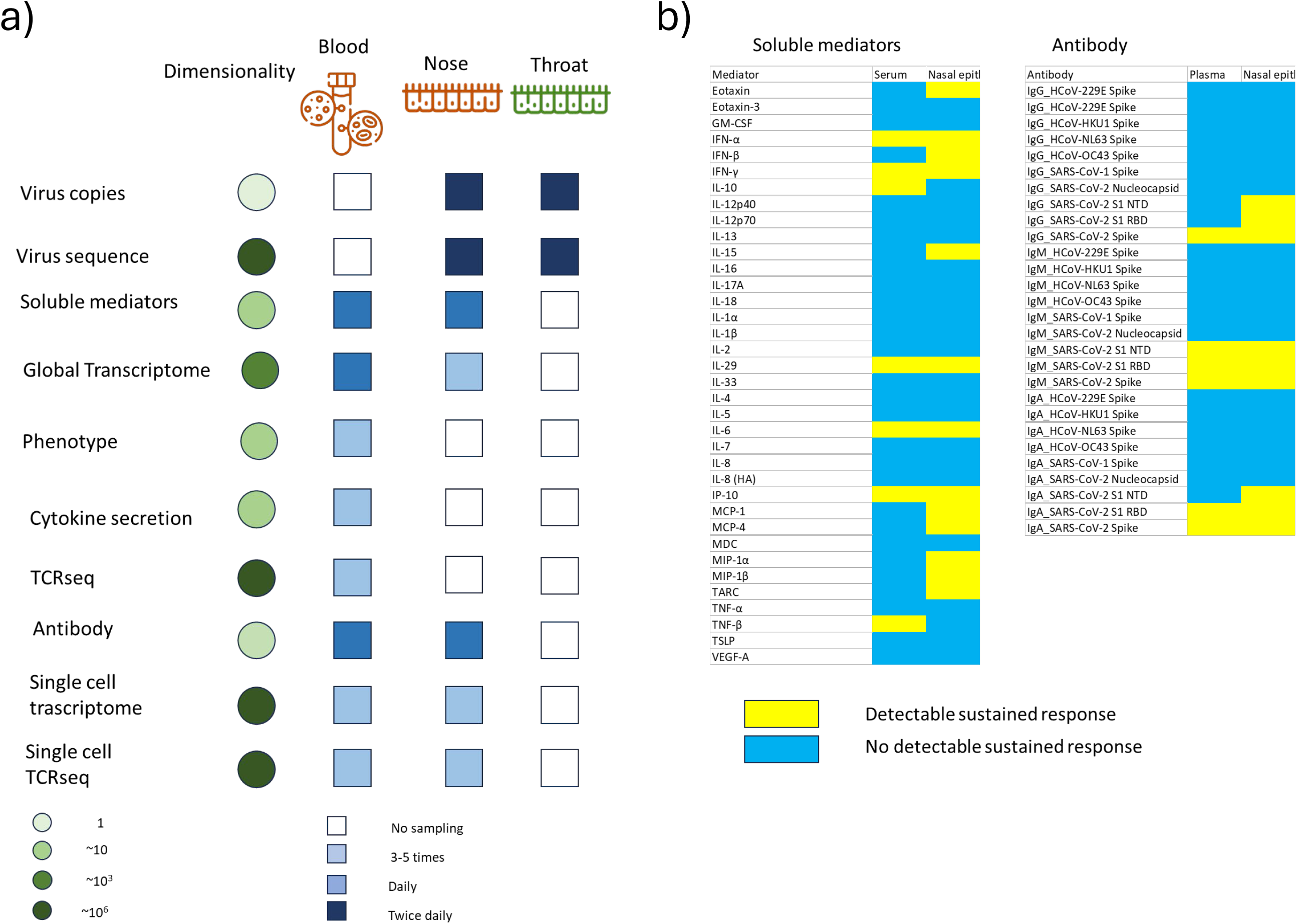
a) A summary of the multimodal data sets collected as part of the first SARS-Cov-2 human challenge study(*15*) b) A list of the soluble mediators measured in serum and in the nasal epithelium as part of the study. Only those mediators detected in a majority of patients in at least one site were analysed further.

**Fig S2.**
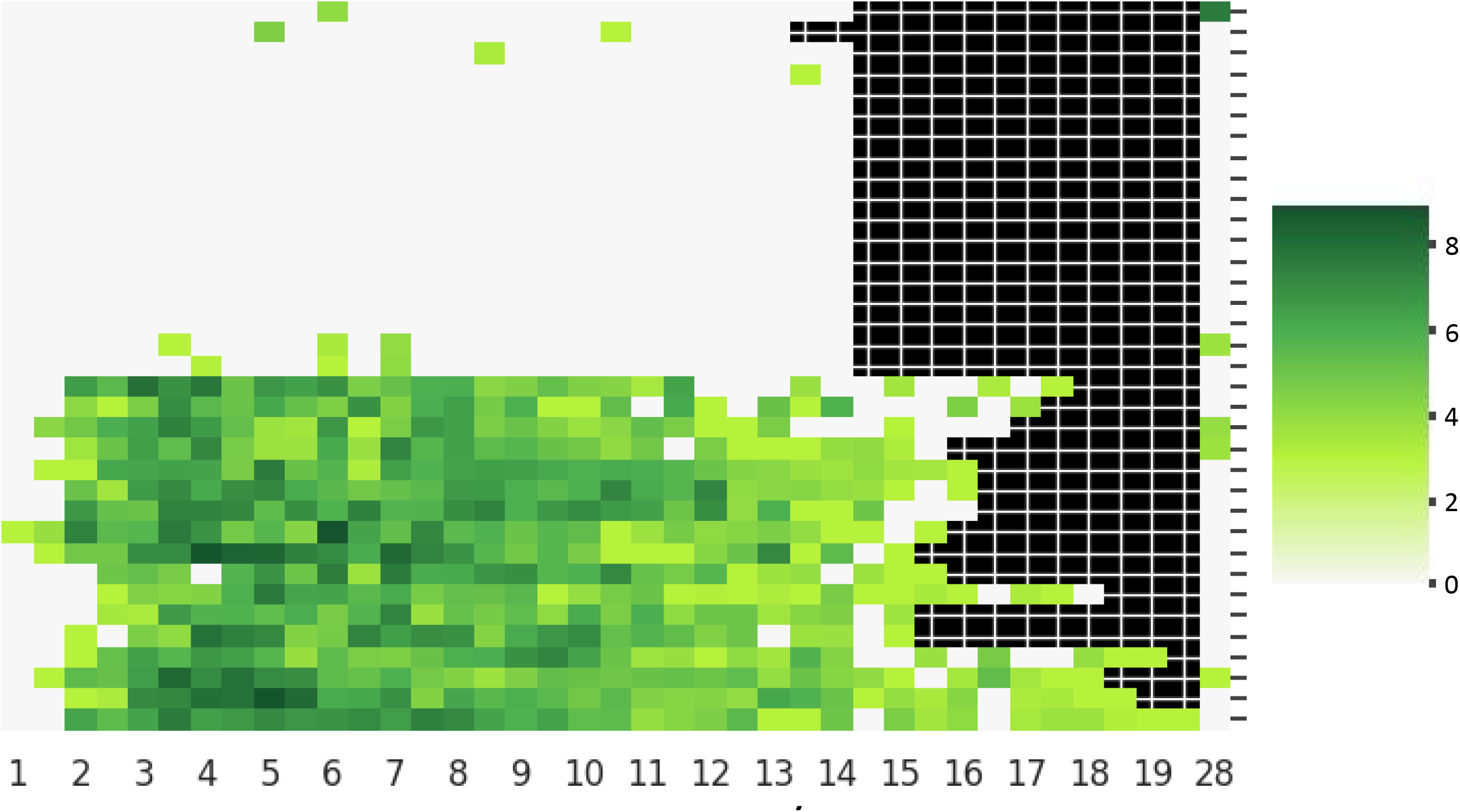
Viral load measured by qPCR (Log10 units) in swab samples taken from the throat mucosal surface. Each row is a different individual. The rows are clustered based on Euclidean distance.

**Fig S3.**
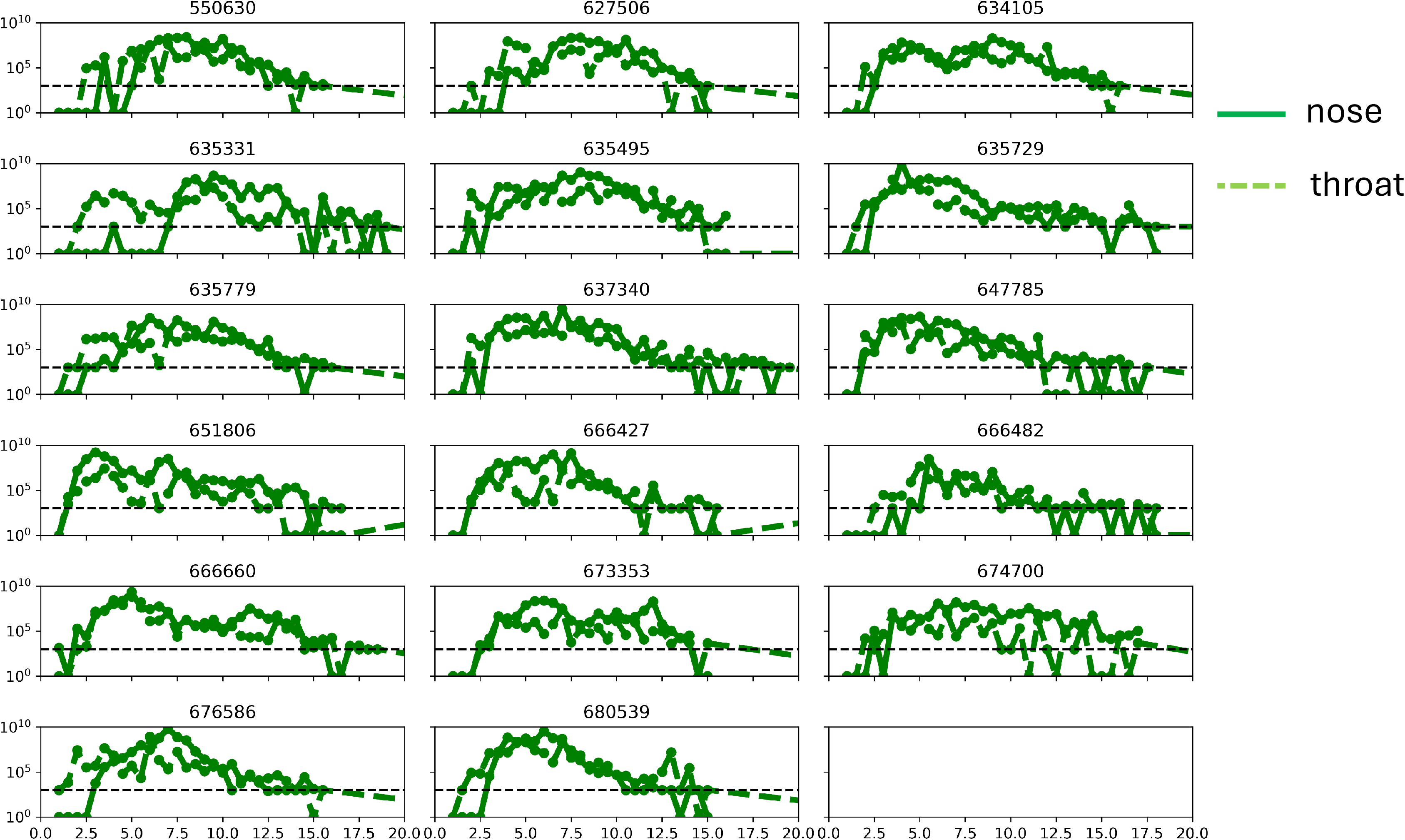
Viral load measurements in nose and throat samples for each individual with a minimum of three consecutive PCR+ samples.

**Fig S4.**
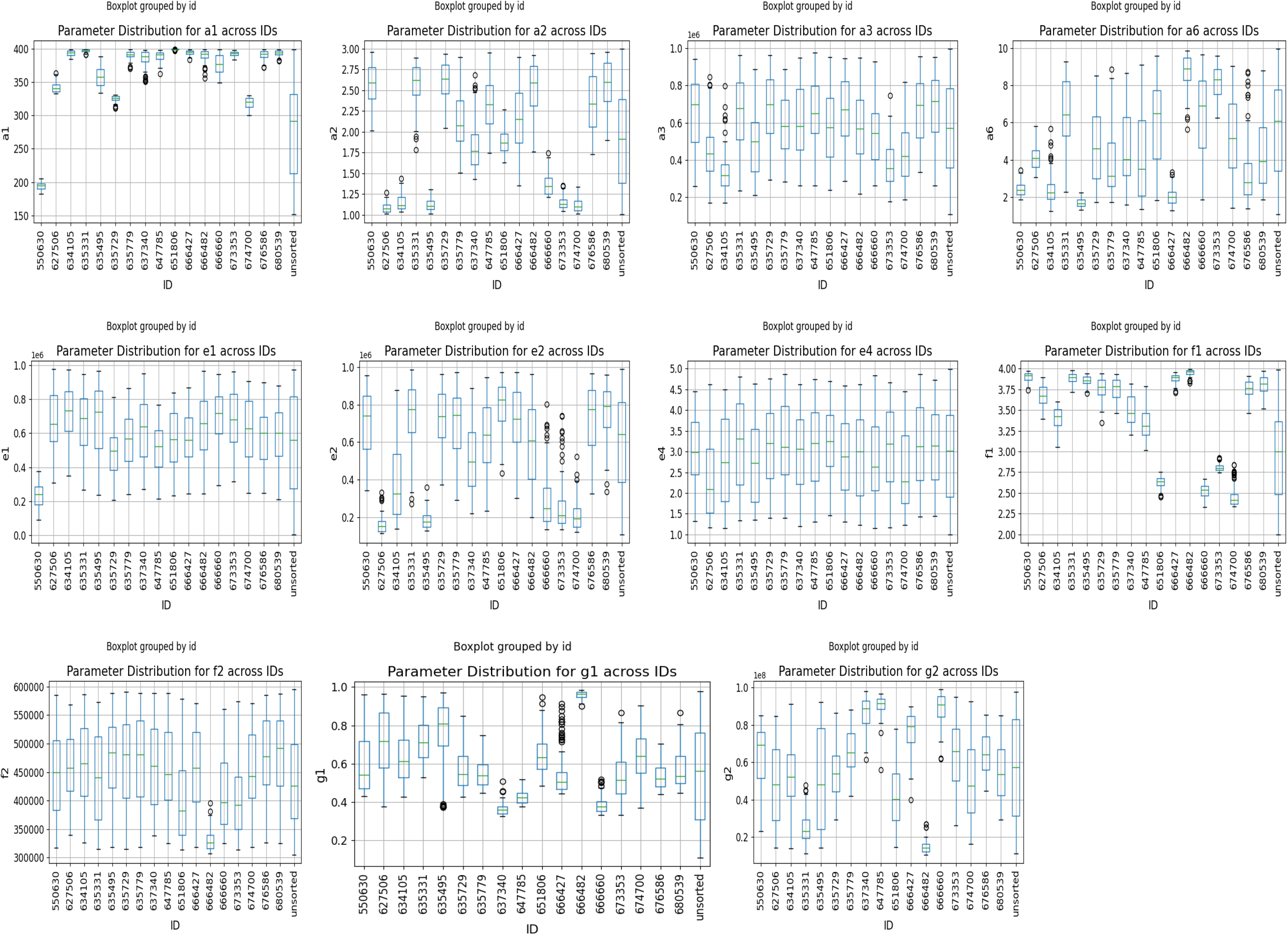
The posterior parameter distributions for the model after Approximate Bayesian Inference. The right hand column in each panel shows the prior distribution of the parameters, randomly and uniformly picked within the limits shown on the y axis. The parameter names correspond to the equations in Fig 1b. Further details are shown in Table S1.

**Fig S5.**
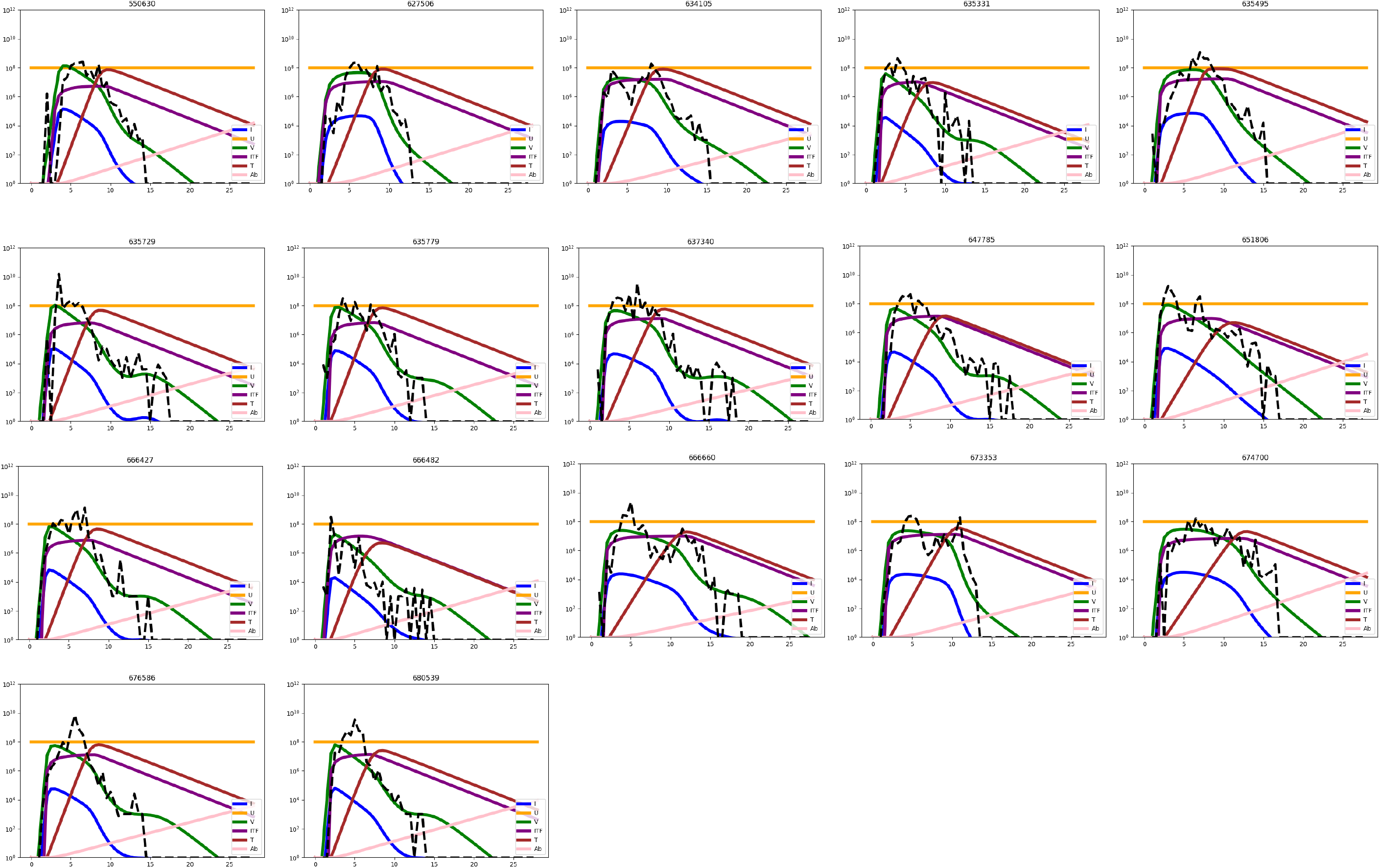
The dynamic profiles of variables shown in the equations in Fig 1b. The plots show the best solution (lowest error) for each individual. Green – virus; Blue – Infected cells; Orange – uninfected cells; Purple – interferon; Brown – Cytotoxic T cells; Pink – antibody. The experimentally measured viral load measurements in the nose, rescaled along time axis, are shown as a dashed black line. The pDC variable is not plotted for clarity, but closely tracks the interferon.

**Fig S6.**
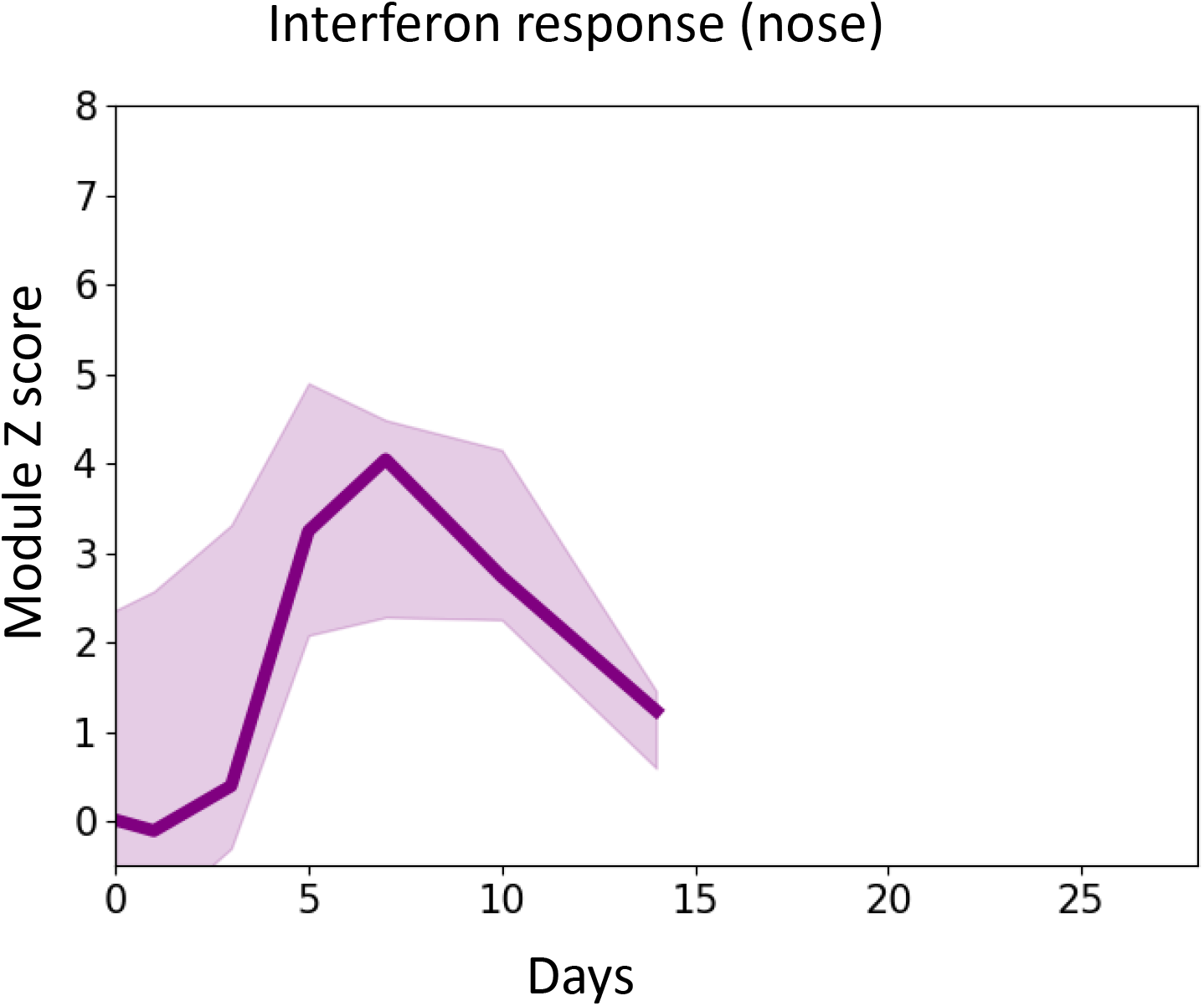
The interferon response transcriptional signal (Z-score Log2 Transcripts per Million median and 95% confidence limits) in the nasal epithelium. No samples were available from the nose after 14 days. The list of genes in the interferon response module are given in Table S2.

**Fig S7.**
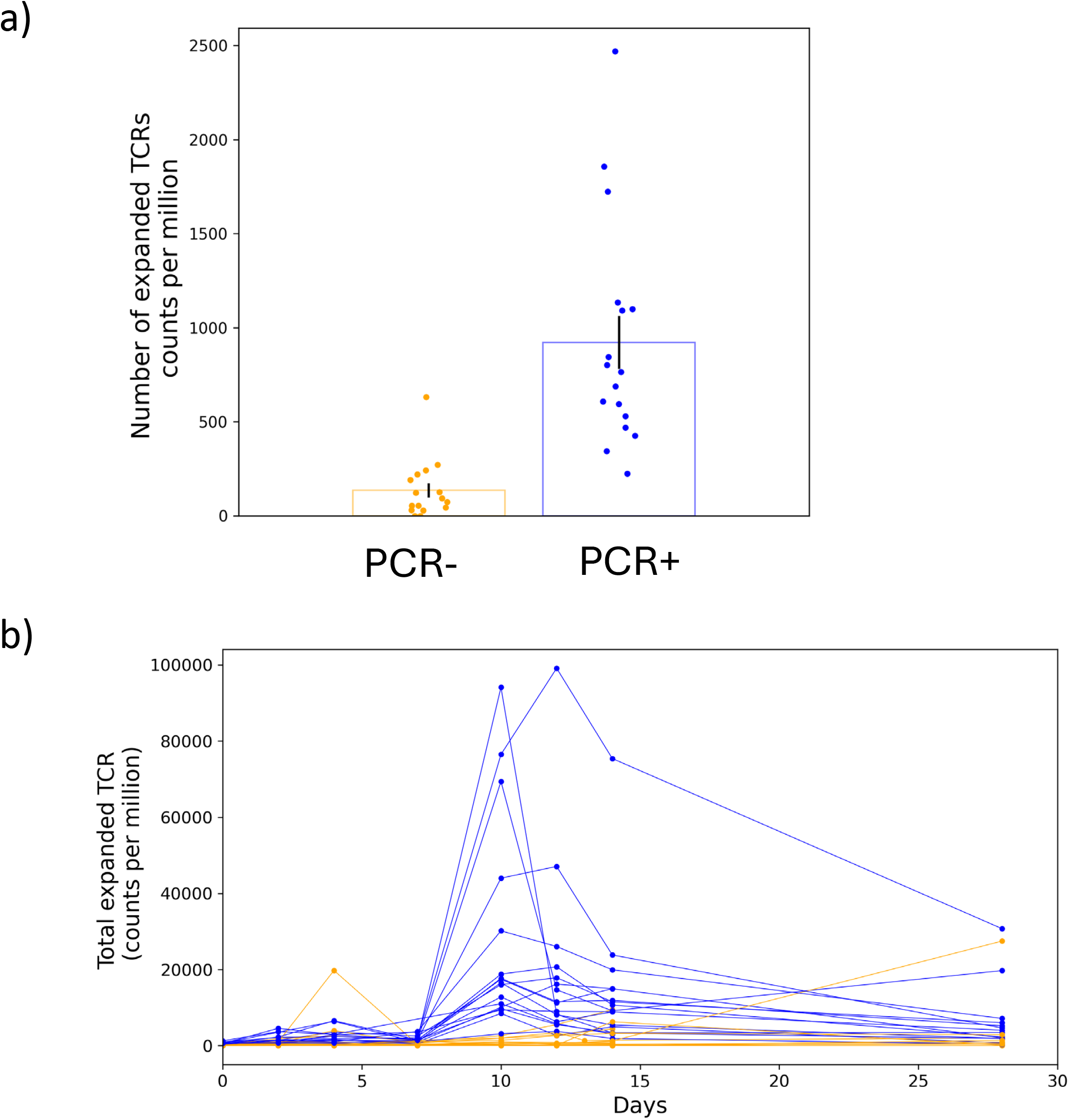
a) The number of expanded TCR β chain sequences in volunteers who do or do not develop a productive infection (cf Fig 2b). b) The sum of the TCR beta clonal frequencies for each individual as a function of time in volunteers who do (brown) or do not (tan) develop a productive infection.

**Fig S8.**
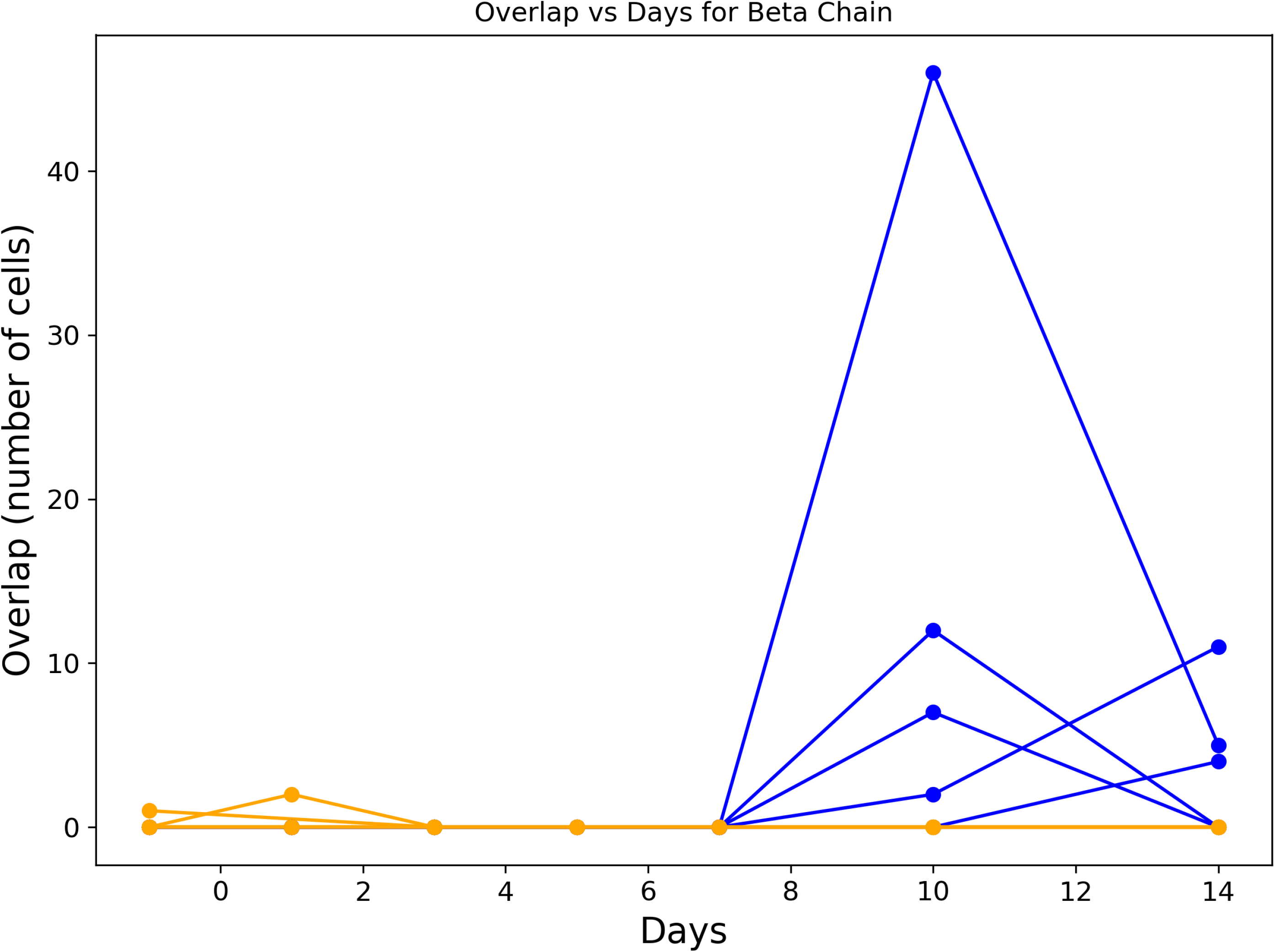
The number of expanded TCRβ chains observed in both blood and nasal epithelium. The TCR sequences from the nasal epithelium were obtained by single cell transcriptomics as described in detail in (*16*).

**Fig S9.**
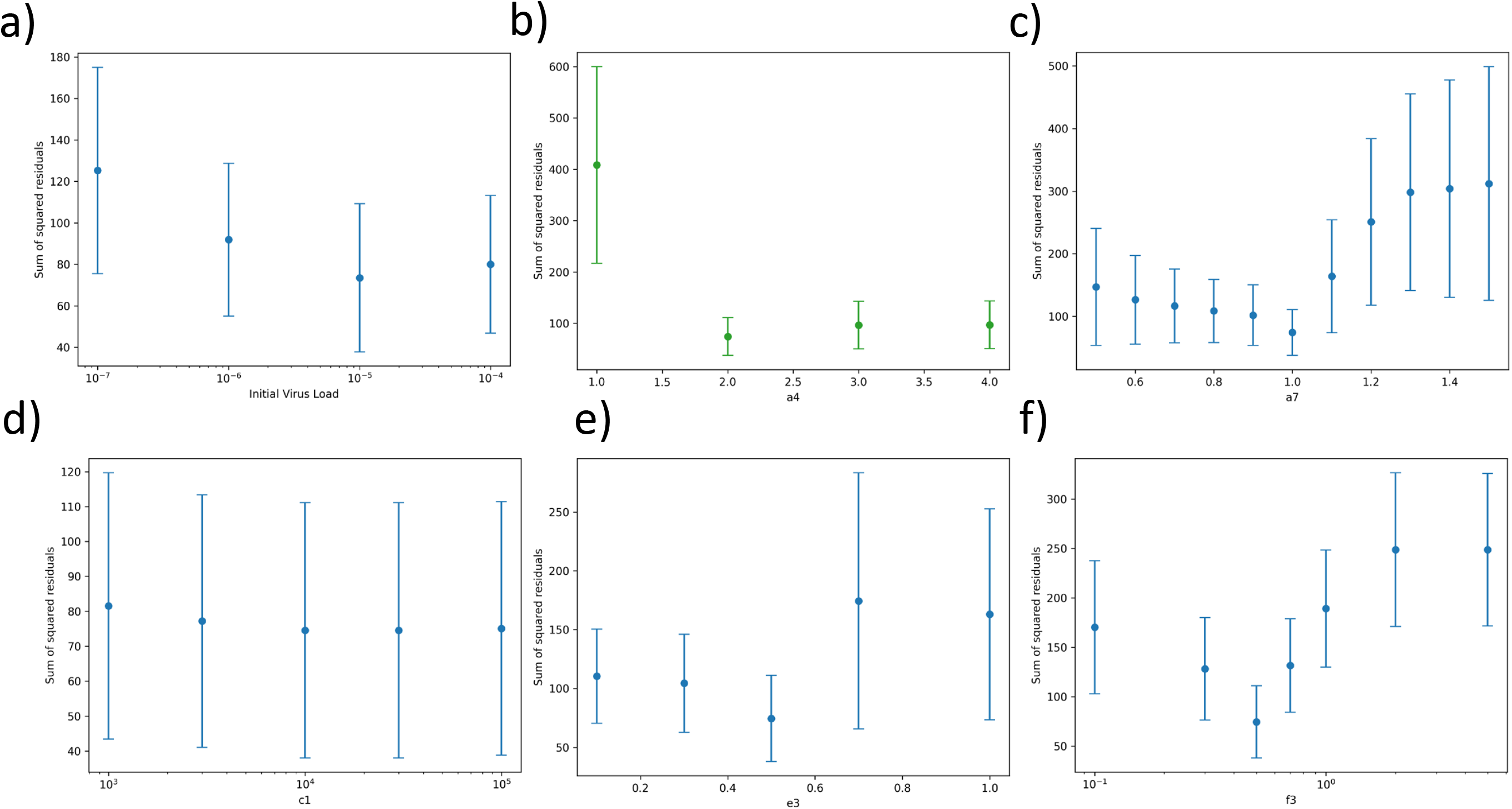
Sensitivity analysis for initial virus load and selected fixed parameters of model as discussed in Methods. The parameter names correspond to the equations in Fig 1b. Further details are shown in Table S1.

**Fig S10.**
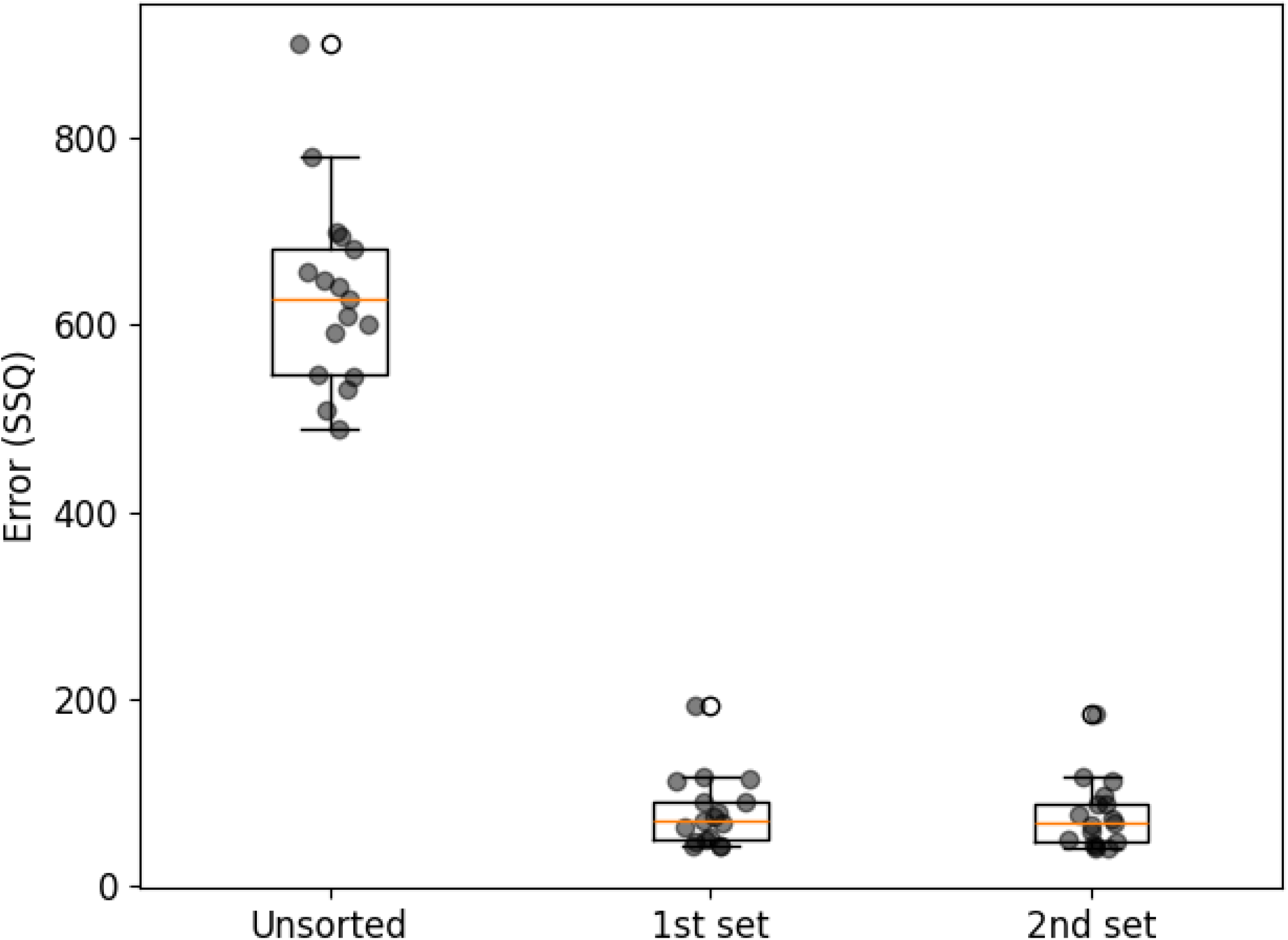
The squared Euclidean distance between the measured viral load and the viral load predicted by the model summed over all sample time points. The unsorted values show the average prior error of 100 model solutions before Approximate Bayesian Inference. The 1^st^ set shows the average posterior error of 100 model solutions with the lowest errors after 10^6^ model solutions. The 2^nd^ set shows the average posterior error of 100 model solutions with the lowest errors after 10^6^ additional model solutions, and restricting the parameter range to the 95 percentiles observed in the top 100 solutions from the first set. The plot shows median and interquartile range. The whiskers extend from the box to the farthest data point lying within 1.5x the inter-quartile range (IQR) from the box. Points past the end of the whiskers are shown individually.

**Fig S11.**
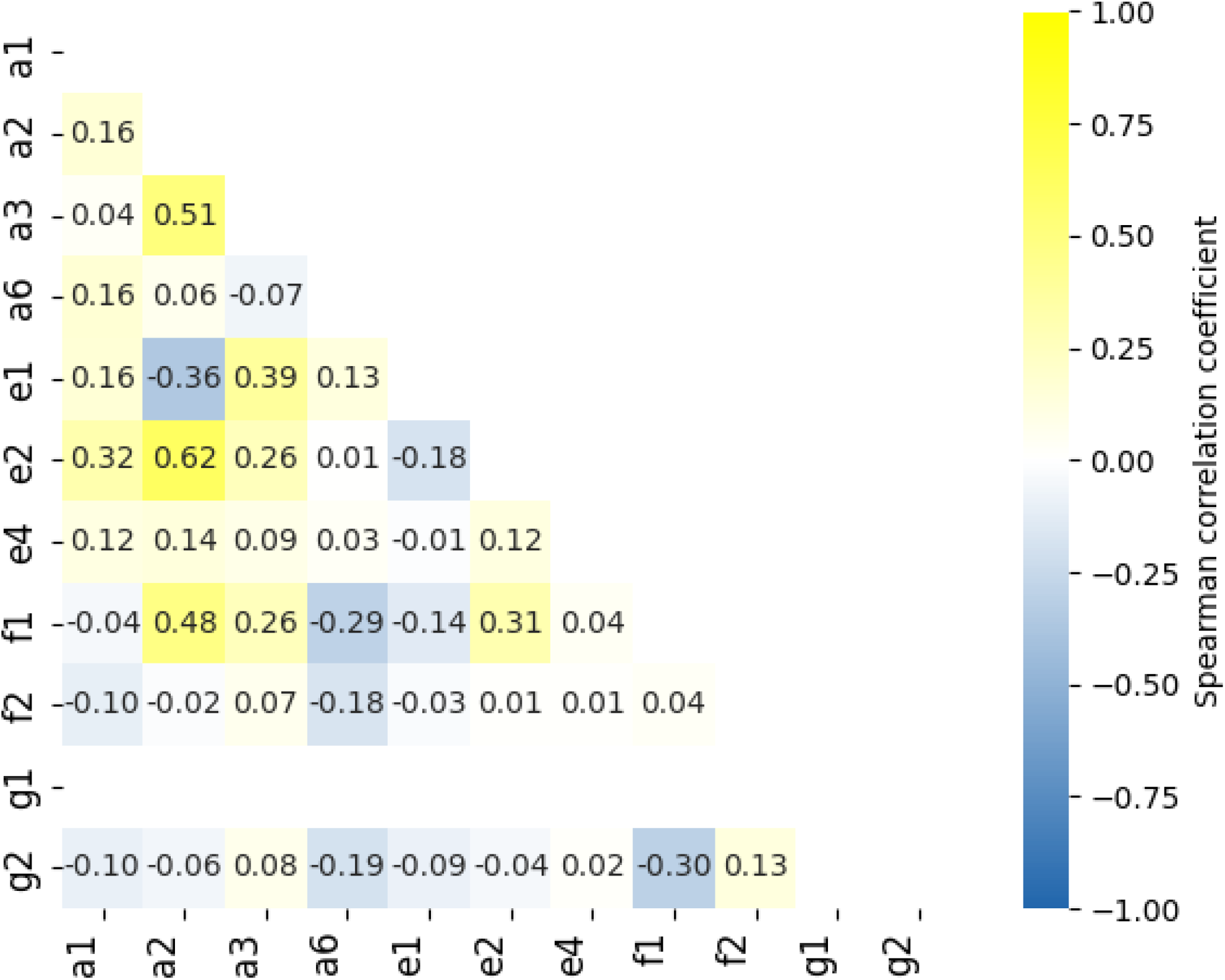
A heat map showing Spearman’s correlation between all pairwise variable parameter combinations. The correlations were calculated for the top 100 model solutions across all individuals.

**Fig S12.**
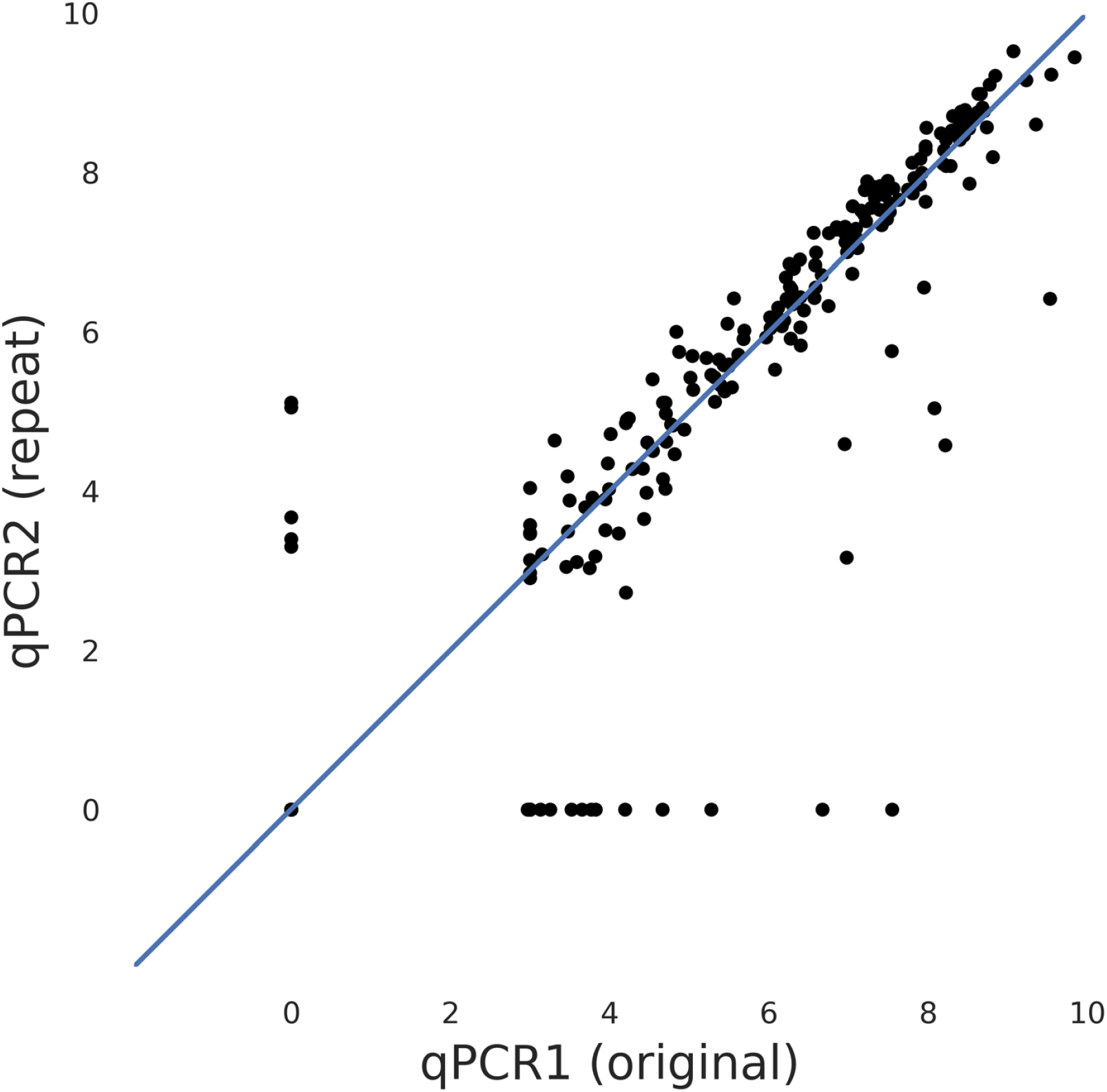
Correlation between replicate viral load correlations made on the same samples by two independent measurements.

**Fig S13.**
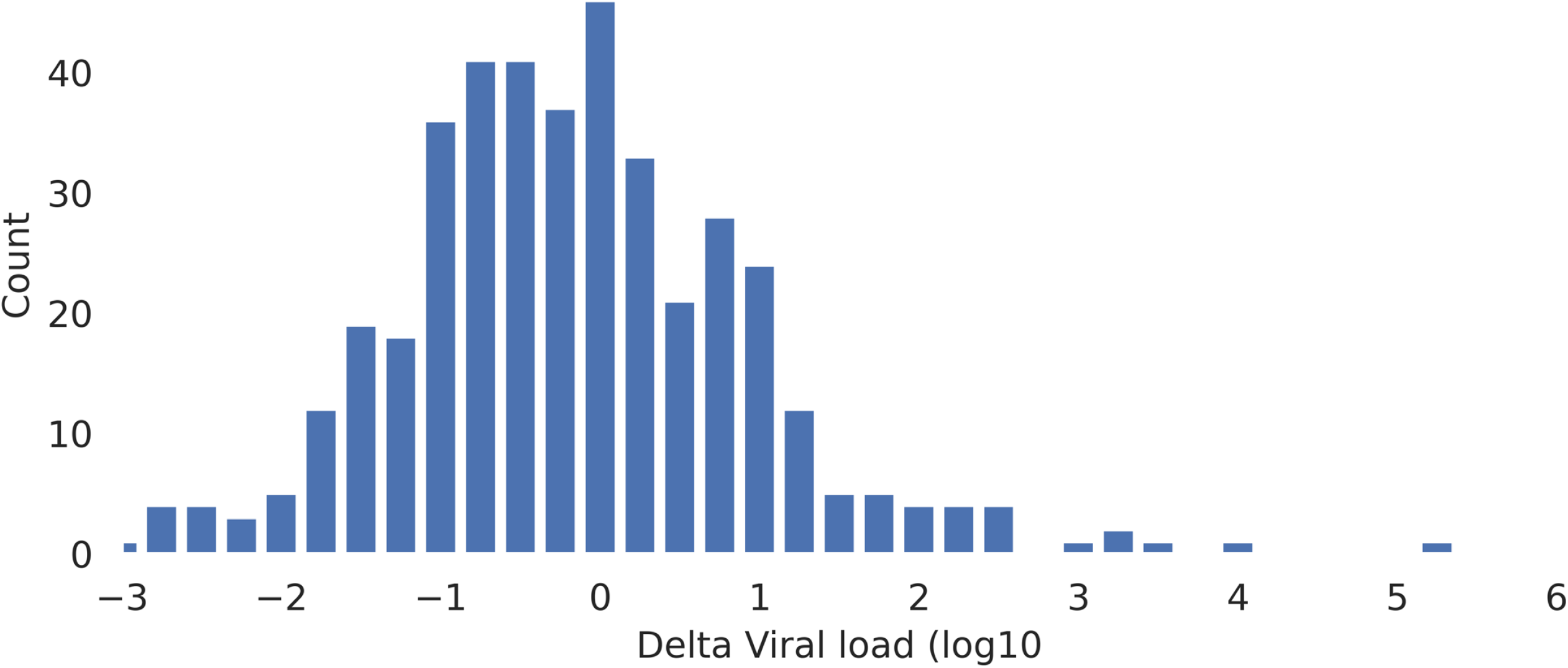
Frequency histogram showing the distribution of differences between viral load measurements made at adjacent time points.

**Table S1.**
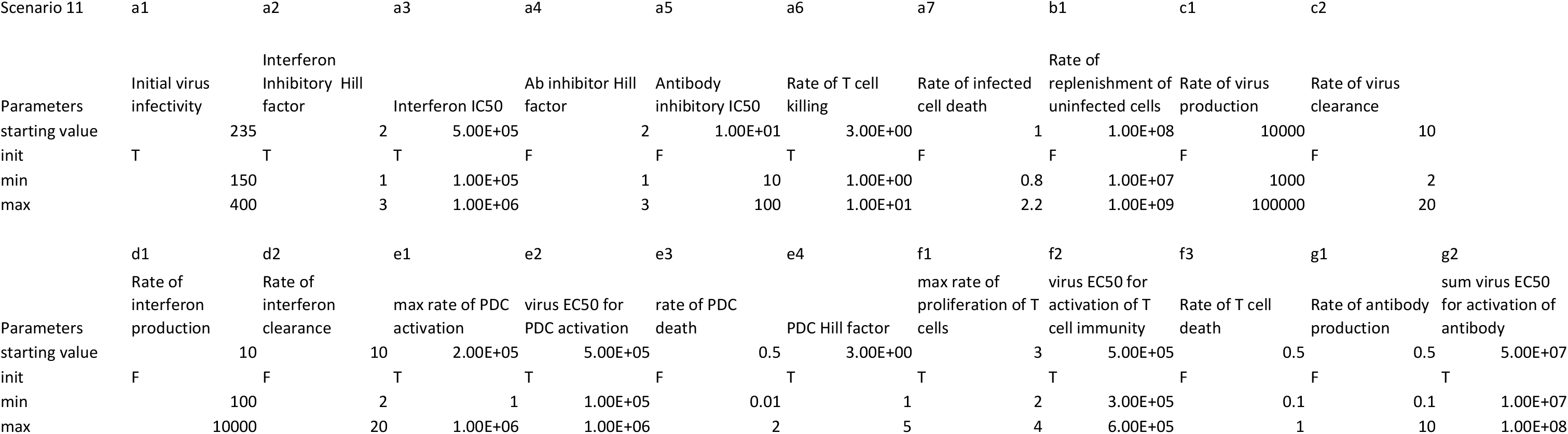
A summary of all the parameters from the equations in Fig 1b. The 2^nd^ row gives a brief summary of the biological interpretation of the parameter. The 3d row column 1 gives the parameter set number. The other columns in row 3 give the value of the parameter for the fixed parameters. The 4^th^ row indicates which parameters are fixed (F) and which parameters are fitted (T). For fitted parameters the values are picked by uniform sampling between the limits shown in rows 5 and 6.

**Table S2.** The list of genes comprising each transcriptional module used in the analysis.

| Short.name | Module.name | Module.genes |
| --- | --- | --- |
| ImmGlob | Immunoglob | IGHD,IGHM,IGHG1,IGHG2,IGHG3,IGHG4,IGHA1,IGHA2,IGHE,IGKC,IGLC1,IGLC2,IGLC3,IGLC7 |
| activeT | Activated.T | CD38,CD28,CD27,ICOS,SELL,GZMK,PRF1 |
| IFN1 | STAT1_Cov | ACOD1,APOBEC3B,BATF2,BIRC5,CCL2,CCNE1,CFB,CMPK2,CXCL10,CXCL11,DDX60,EIF2AK2,EPSTI1,HERC6,IFI27,IFI44,IFI44L,IFI6,IFIH1,IFIT1,IFIT3,IFITM3,IGF1,IL10,IL21,IRF7,ISG15,LY6E,MX1,OAS1,OAS2,OAS3,OASL,RSAD2, RTP4,SAMD9L,SERPING1,STAT1,USP18,XAF1,ZBP1 |
| Prolif | CCND1_Co | ASPM,AURKA,BIRC5,CCNA2,CCNE1,CDC45,CDC6,CDCA2,CDCA7,CDCA8,CENPF,CENPN,CEP55,CIP2A,CLSPN,DEPDC1,DHCR24,DTL,E2F1,E2F7,E2F8,ESCO2,FAM83D,FOXM1,H4C8,HJURP,ID1,KIF11,KIF20A,KIF20B,KIF2C,KIF4A,KNL1,KRT14,MCM10,MCM4,MELK,MTFR2,NCAPH,PCLAF,PCNA,PLK1,RAD51,RRM2,SPC25,TMPRSS2,TMSB15A,TPX2,TRIP13,TYMS,UHRF1 |

## Author contributions

Conceptualization: BC, AT-M, CC, DS, MN

Data acquisition: JR, CT, LB, GN, SB, BK, RGHL

Data curation and analysis: BC, CCa, DS, JJ, RG, MVC, CTT, AT-M, NM, RGHL, HW

Funding acquisition: BC, CCh, MN, SAT

Resources: MK, AB, AC, AM, MZN, MN, CCh

Writing – original draft: BC, MN

Writing – review & editing: All authors

## Data and materials availability

### Materials availability

This study did not generate new unique reagents.

### Data and code availability

The SARS-Cov-2 Challenge TCR sequencing data have been deposited at NCBI Sequence Read Archive (https://www.ncbi.nlm.nih.gov/sra) and are publicly available under project accession number ??. The processed T cell receptor sequence files, and all other data sets and files used in this study are deposited at 10.5281/zenodo.22832153. All code for analysis and generating individual figure panels are available on GitHub https://github.com/innate2adaptive/Modelling-SARS-COV-2-challenge/ The analysis was carried out using Python v3.10.12. Any additional information required to reanalyse the data reported in this paper is available from the lead contact upon request. Requests will be responded to within 4 weeks of receipt.

